# Association of Long-Term Outdoor Air Pollution Exposure with Incidence of Parkinson’s Disease, Multiple Sclerosis and Motor Neuron Diseases: A Systematic Review and Meta-Analysis

**DOI:** 10.1101/2025.04.15.25325911

**Authors:** Alexandra Z. Tien-Smith, Shazia Absar, Clare Best Rogowski, Veronica Phillips, Zorana Jovanovic Andersen, Christiaan Bredell, Kwan Wai Fung, Lucy Hong, Magdalena Szybka, Stephen Sharp, James Woodcock, Carol Brayne, Haneen Khreis, Annalan M D Navaratnam

## Abstract

**Background:** Parkinson’s disease (PD), multiple sclerosis (MS) and motor neurone disease (MND) are progressive and debilitating diseases that are increasing in prevalence globally. Some primary studies show an increased risk from long-term outdoor air pollution exposure, while others contradict this association.

**Methods:** As per Khreis et al.^1^ protocol, a systematic review and meta-analysis was undertaken to assess the associations of long-term (≥1 year) outdoor air pollution exposure with PD, MS and MND incidence. We searched eight databases for publications up to October 2023. Primary case-control, cohort, cross-sectional or ecological studies investigating the association of long-term air pollution and adult (>18 years old) PD, MS, or MND incidence were included. Meta-analyses were carried out using random-effects models with assessment of heterogeneity and meta-bias. PROSPERO (CRD42023417961).

**Results:** Of 31 papers included, 22 and 3 were meta-analysed for PD and MS outcomes, respectively. Most studies were from North America (14) followed by Europe (8), and Asia (6). For every 5 μg/m^3^ increase of Particulate Matter 2.5 (PM_2.5_) concentration, there was a higher PD risk (1.06; 95%CI: 1.00-1.12), but this was not true for all study settings (Prediction Interval: 0.95-1.19). This risk was largest in Asia (1.16, 95%CI:0.96-1.41). There was no evidence that PM_2.5_ or nitrogen dioxide (NO_2_) were associated with increased risk of MS.

**Conclusion:** This systematic review reports an increased risk of PD from long-term PM_2.5_ exposure. The neurodegenerative diseases investigated here are rare and therefore alternatives to insufficiently powered cohort studies are needed to strengthen the evidence on risk.

## Background

Outdoor air pollution is of critical public health concern and is a major contributor to the global burden of disease.^2^ Fine particulate matter (PM_2.5_), nitrogen dioxide (NO_2_) and ozone (O_3_), have been associated with an increased risk of various neurological diseases, including stroke and dementia^3,4^. They have also been implicated in neurodegenerative diseases such as Parkinson’s Disease (PD), Multiple Sclerosis (MS) and Motor Neurone Disease (MND), which are characterised by progressive and debilitating impairments in motor function, significant deterioration in quality of life and premature mortality.^5–7^ At present, these conditions are not curable and treatments cannot reverse progression. Therefore, it is essential to understand the roles that modifiable risk factors, such as outdoor air pollution, play to tackle these growing public health challenges. ^8^

PD is the second most common neurodegenerative disorder worldwide, affecting approximately 6 million people. ^9^ It predominantly occurs in the elderly and has substantial morbidity and disability.^9^ Deaths, Disability Adjusted Life Years and prevalence have increased between 1990 and 2016, even when adjusting for the ageing population.^10^ MS, a chronic autoimmune condition that affects the central nervous system, has also increased in prevalence, with an estimated 2.8 million cases worldwide.^10,11^ There have been changes in demographic (e.g., sex and ethnicity) and geographic distribution of MS disease burden overtime, which may reflect changes in environmental factors as drivers of susceptibility.^10,12^ MND is less common but equally devastating, causing severe disability with a high fatality rate.^13^ Amyotrophic Lateral Sclerosis (ALS), the most common MND, causes respiratory failure in 50% of patients within 2 years of diagnosis. ^14^ The burden of MND has increased substantially with increases in population ageing and growth.^10,15^ The rising global burden of these diseases presents significant public health challenges, both in terms of healthcare costs and quality of life.

While the aetiology of PD, MS, and MND is multifactorial, growing evidence suggests that environmental factors, including outdoor air pollution, may play a pivotal role in their onset and progression.^1^ PM_2.5_, which penetrates deep into the lungs and circulates through the bloodstream, has been shown to contribute to neuroinflammation, oxidative stress and mitochondrial dysfunction, mechanisms that are central to the pathogenesis of these neurodegenerative diseases.^16–20^ Studies linking exposure to air pollution with PD have highlighted potential mechanisms through which pollutants may affect dopaminergic neurons.^21,22^ The relationship between air pollution and MS remains less well understood but is thought to involve immune dysregulation.^23^ Although there is a scarcity of evidence on risk factors for MND, some research implicates exposure to higher concentrations of PM_2.5 absorbance_ and NO_2_ are associated with higher risk of ALS compared to lower concentrations.^15,24^

Given the increasing global prevalence of these diseases and the growing body of literature on the health effects of air pollution, it is critical to systematically review the evidence to clarify the extent to which outdoor air pollution may contribute to the risk of these diseases. Several meta-analyses of epidemiological studies on the association between outdoor air pollution and PD have been published.^25–30^ There are inconsistent conclusions between these meta-analyses however, due to insufficient number of primary studies, marginal pooled effect estimates, high inconsistency and high heterogeneity.^1^ Among the six published meta-analyses identified in the literature, only two demonstrated a statistically significant association of PM_2.5_ exposure with PD risk, but with widely varying effect estimates (1.34 per 10 μg/m^3^ increase; 95% Confidence Intervals [CI]: 1.04-1.73 with seven studies up to the year 2018 and 1.01, 95% CI: 1.01-1.02 with eight studies up to the year 2018, respectively).^26,29^

For gaseous pollutants, evidence for an association of oxides of nitrogen (NO_x_) and NO_2_ using pooled estimates of relative risk for PD has been conflicting. Hu et al^28^ identified an association of NO_x_ exposure (1.06 per 10 ppb increment; 95% CI: 1.04- 1.09 with four studies up to the year 2017) whereas Han et al^27^ reported no increase from NO_x_ exposure (1.00 per 1 ppb increment; 95% CI: 0.98-1.03 with four studies up to the year 2019).

To date there are two published meta-analyses that demonstrated an association of outdoor air pollution with increased MS incidence. One, however with studies up to the year 2019, grouped MS incidence and relapse into a single outcome and estimated an increased pooled rate ratio with PM_2.5_ (1.18, 95% CI: 1.10 – 1.28) incidence with four studies and PM_10_ (1.34, 95% CI: 1.13 –1.58) incidence with 10 studies.^31^ The other examined MS incidence alone and found an association using a pooled hazard ratio with PM_10_ with four studies up to the year 2020 (1.06 per 10 μg/m^3^ increase, 95% CI: 1.05 – 1.07).^32^ The latter study also showed increased pooled risk estimates for O_3_, NO_2_ and PM_2.5-10_ exposure, but these did not reach statistical significance.^32^ Lotfi et al. and Tang et al. both reported a small number of included studies as a shortcoming in their investigation of the association of long-term air pollution. Finally, there are no systematic reviews or meta-analyses on MND incidence, and only limited primary research into, the possible association of air pollution with MND.^1^

Since 2022, a rapid increase in publications has provided an opportunity for a more comprehensive systematic review and meta-analysis. It is essential to quantitatively synthesise and interpret the most up to date evidence to provide more comprehensive and consistent information for policy making and further research. This study therefore will examine whether long-term exposure to ambient air pollution increases the risk of adult incidence of PD, MS and MND.

## Methods

A protocol for this systematic review was published in December 2022 and registered with the International Prospective Register of Systematic Review (PROSPERO CRD42023417961).^1^ The deviations from this protocol are listed in Table S1 and are mostly minor and more inclusive in nature.

### Search Strategy

Medline (via Ovid), Embase (via Ovid), Cochrane Library, Cinahl (via Ebscohost), Global Health (via Ebscohost), PsycINFO (via Ebscohost), Scopus, and Web of Science (Core Collection) were searched twice (once from inception to October 2022 and again from October 2022 to October 2023) using peer-reviewed and piloted search terms developed by the authors through a review of existing literature, a selection of indicator papers, and pilot searches, according to the published protocol.^1^ These terms are included in Table S2.

### Eligibility Criteria

Studies were eligible if they were based on the results of a primary case-control, cohort, cross-sectional, or ecological studies of individuals over 18 years of age, investigated exposure to outdoor air pollution of a year or more (long-term), and quantitatively reported the association between air pollution exposure and a subsequent clinical diagnosis of PD, MS or MND in adults free of disease at baseline. We excluded papers that exclusively reported on markers or prognosis markers of the health outcomes of interest and not the health outcomes specifically. We also excluded studies that did not provide a quantitative exposure estimate or used a proximity model for exposure assessment. Complete inclusion and exclusion criteria are in Table 1.

**Table 1.** Inclusion and exclusion criteria [Khreis 2022].

| Inclusion criteria | Exclusion criteria |
| --- | --- |
| English language.<br>Peer reviewed and published in a journal.<br>Either case-control, cohort, cross-sectional, or ecological. For cross sectional studies the design must be sufficient to answer the primary research question reported as incidence or life-time prevalence.<br>Study population must consist exclusively of adults or analyse adults alone as a subgroup.<br>It must be a primary study that investigates the effect of long-term ($\geq 1$ year) exposure to outdoor air pollution, on PD, MS, and MND. Diagnosis must come from either a clinical diagnosis (medical, prescription, or insurance records) or a self-report of an existing clinician diagnosis. Self-diagnosis is not acceptable.<br>The association between preceding exposure to air pollutants and the outcomes of PD, MS, and MND are reported quantitatively [i.e., Incidence (life-time prevalence acceptable only in cross-sectional studies), or risk estimates, including odds ratios (OR), hazard ratios (HR) or relative risk ratios (RR)]. The comparison must be between those who were exposed to more air pollution and those exposed to less air pollution. | Study population is not adult humans (or does not include them as a subgroup).<br>Exposure was prenatal.<br>Study only investigated exposures other than outdoor air pollution.<br>Diagnosis was ascertained by self-diagnosis.<br>Study design is not case-control, cohort, cross-sectional, or ecological (i.e., All non-epidemiological study designs, a short term-study which may include case-crossover studies, time-series studies).<br>Does not report a quantitative risk estimate for the association of PD, MS, and MND with air pollution.<br>Not peer-reviewed, or is a conference abstract, preprint, or thesis.<br>Not a primary study (i.e., A literature review, systematic review, or meta-analysis).<br>Reports exclusively on exacerbations of PD, MS, and MND (i.e., Hospitalisations) when it pre-existed, and not incidence or lifetime prevalence.<br>Reports on prognosis markers of the health outcomes of interest exclusively (i.e., Does not report on PD, MS, and MND outcomes specifically).<br>Used a proximity model for exposure assessment, which does not collect a numerical exposure variable and has higher risk of exposure misclassification and residual confounding (Jerrett et al., 2005, Zou et al., 2009) |

### Selection Process

Deduplication of the search results was completed in EndNote then Zotero and then Rayyan. The remaining papers were title and abstract screened by two independent reviewers. Papers included by at least one reviewer underwent full-text screening by two reviewers; any remaining conflict was resolved by a third reviewer. The list of studies excluded at both the title and abstract and full text screening stages, alongside the reason for exclusion are in Supplementary Item (SI) 1. The original search included dementia outcomes, the topic of another paper currently under review, alongside PD, MS and MND (including names for specific diseases within this group as shown in Table S2).^1,3^ Following title and abstract screening, papers were divided by outcome for separate data extraction and analysis.

### Data Extraction

Using a standardised and piloted form through multiple iterations (SI 2), data was extracted independently from each included study by two reviewers.^33^ Duplicate sheets were compared, and discrepancies were resolved. Unadjusted and adjusted measures of association (effect estimates) were recorded with their reported 95% CI, unit of exposure (µg/m^3^, ppb, etc), scaling factor (e.g., 1 µg/m^3^, 5 µg/m^3^, 10 µg/m^3^), and covariates adjustment. Additional data on, for example, study type, location, year, length of follow-up, population, exposure assessment, outcome assessment, and funding were also recorded (Table S3). The complete list of extracted data points is included in SI 3. If one paper reported more than one effect estimate per pollutant, the estimate that met the criteria in the published protocol (Table 2) was extracted.^1^ If information could not be determined for a paper, we attempted to contact the authors to clarify, and if no response was received after three attempts it was coded as ‘Not Available’.

**Table 2.** Effect Estimate Selection Criteria.

|  |  |
| --- | --- |
| Where more than one effect estimate per pollutant (exposure-outcome pair) is reported in one study, we selected for inclusion the risk estimates that: |  |
| 1. | Related to the longest or most cumulative exposure window during the follow-up period with the assumption that this is the most relevant biological window. |
| 2. | Was most inclusive in capturing the outcome over the follow-up (e.g., incidence over 10 years versus 5 years) or that which emerged from the most recent follow-up. |
| 3. | Related to the most definitive or objective outcome definition (e.g., a clinician's diagnosis versus a self-report of a clinician's diagnosis). |
| 4. | Related to models including single pollutants (risk estimates from multi-pollutant models will be recorded separately |
| 5. | Related to the total population in the wider geographical area rather than regional or sub-geographical estimates. |
| 6. | Was estimated using the exposure model with the highest spatial resolution (e.g., land use regression model versus fixed site estimates). |
| 7. | Related to the most restrictive analysis model (e.g., including adjustment for lifestyle factors such as smoking, alcohol consumption, diet, and physical activity, indoor environmental factors, versus not), if we judge the adjustment as appropriate. Whether the adjustment is appropriate was determined through our risk of bias assessment (e.g., not adjusting for co-morbidities on the pathway between air pollution and PD, MS, and MND outcomes). This is particularly relevant for neurodegenerative diseases where there is evidence that diseases such as stroke, heart failure, and ischemic heart disease, can be intermediate conditions and/or effects modifiers and explain a significant proportion of air pollution-related PD, MS, and MND cases. Diabetes and depression may also be intermediate conditions and/or effect modifiers. Adjustments for these factors may be inappropriate. If adjusted effect estimates are only available from models which include these morbidities, we will include these estimates in the meta-analysis but will conduct a sensitivity analysis to describe the impact of removing them. |

### Risk of Bias Assessment

Using the Office of Health Assessment and Translation (OHAT) tool^34^, two independent reviewers assessed Risk of Bias (RoB) for each included study (internal validity), with modifications implemented after piloting the tool at the outset (SI 6). ^1^ While the OHAT tool includes 11 questions, 4 of these questions were excluded, because they are related to Experimental Animal and Human Controlled Trials, making them inapplicable. For each of the seven remaining questions, one of the following four answers was selected: definitely low risk of bias, probably low risk of bias, probably high risk of bias, definitely high risk of bias. Discrepancies between reviewers were discussed and a consensus was reached, and if conflicts emerged, they were escalated to a third reviewer.

### Certainty of Evidence Assessment

Two reviewers rated the confidence in evidence bodies using the OHAT approach for systematic review and evidence integration for literature-based health assessments.^35^ Each article was ranked as “high confidence,” “moderate confidence,” “low confidence,” or “very low confidence.” Because observational studies do not have “controlled exposure,” one of the four key features in rating certainty of evidence, the highest rating the papers could attain was “moderate confidence” provided they had the other three categories: “exposure prior to outcome,” “individual outcome data,” and “comparison group used.” Overall certainty of evidence was then reviewed for factors that decreased (e.g., risk of bias, unexplained inconsistency, imprecision) and increased (e.g., large magnitude of effect, dose-response relationships) confidence. Any discrepancies between reviewers were discussed and a consensus was reached.

## Statistical Analysis

### Standardisation

Following data extraction, all papers were combined for statistical analysis. Some included studies used the same study population, so only the study with the longest follow-up in meta-analyses was included to avoid double counting. Meta-analyses were conducted when three or more independent studies reported an exposure-outcome pair (e.g., PM_2.5_ and PD, PM_2.5_ and MS, or PM_2.5_ and MND). For each pollutant, adjusted and unadjusted effect estimates, and their 95% CI were harmonised to a prespecified exposure increment representing 2021 World Health Organization Standards. Annual WHO concentration guidelines were not available for PM_2.5-10_, NO_x_, O_3_ (annual and warm season), CO, and SO_2_, so concentration increments were standardised to 10% of the maximum concentration reported in any included study. If NO_2_, NO_x_, O_3_ (annual and warm season), CO, and SO_2_ were reported as parts per million (ppm) or parts per billion (ppb), they were converted μg/m^3^, with the exception of CO which was converted into the standardised unit mg/m^3^.

### Meta-Analysis

The DerSimonian and Laird (DSL) inverse-variance method, the most widely employed, was used to weight the harmonised effects estimates for directness of comparison with other meta-analyses.^36,37^ When outcomes are rare (i.e., <10-20%), the hazard ratio (HR) and odds ratio (OR) can be approximated to a risk ratio (RR).^38^ Given that neurodegenerative movement disorders are considered a rare outcome, pooling the HR and OR for each outcome together in one meta-analysis in this case was considered acceptable. The analysis of unadjusted effect estimates was designed to give insight into the effects of (residual) confounding factors.

### Measuring Heterogeneity

Between-study inconsistency and heterogeneity were quantified using the I^2^ statistic and Cochran’s Q-test (Q), to determine whether differences between study results account better for the variability in the meta-analysis than sampling error, in addition to prediction intervals (PIs).^39–41^ One of these tests alone is not sufficient: the value of I^2^ heavily depends on the precision of the included studies and Q is known to have low statistical power in small meta-analyses. These three measures together provide a more meaningful comment on whether the observed differences in the results of the meta-analysis may reflect true effects rather than sampling error.^42^

### Subgroup and Sensitivity Analyses

The published protocol outlined that we would conduct subgroup analyses by geographical location, sex, age, ethnicity, exposure assessment method, outcome definition and ascertainment method.^1^ The number of studies with data for sex, age, ethnicity, and outcome definition and ascertainment method were insufficient (i.e., fewer than three studies). Subgroup meta-analyses were therefore only undertaken on geographical region, exposure assessment method, and effect estimate for PD.

As per the protocol, we conducted the following sensitivity analyses: (1) excluding the study with the largest weight (smallest SE), (2) excluding the studies with a high risk of bias, which are defined as studies with at least one domain with a “definitely high risk of bias” rating, (3) running the analysis separately for studies with “Definitely low risk of bias” and “Probably low risk of bias” combined, versus “Probably high risk of bias” and “Definitely high risk of bias” combined, (4) running the analysis by study design (separately with cohort, case-control, cross-sectional and ecological studies when three or more independent studies reporting an exposure-outcome pair are available), and (5) removing studies that adjusted for any co-morbidities and cardiometabolic comorbidities (e.g., cardiovascular disease, diabetes mellitus). It was not possible to carry out a sensitivity analysis excluding studies that relied on patient self-report of clinical diagnosis for outcome assessment, as all included studies had some form of physician or medical record corroboration. Sensitivity analyses were conducted using the Paule-Mandel in place of the DSL, given reports that the DSL is negatively biased in scenarios with small studies and with a rare binary outcome.^43–45^

Assessing the shape of the Exposure Response Functions (ERF)

To evaluate the shape of a pollutant’s exposure-response function (ERF), we graphed exposure-outcome pairs when four or more independent studies reported on the same exposure-outcome relationship across different exposure levels. Following the protocol, we treated the median of the upper and lower values of each exposure category as representative of that category’s exposure level when establishing these pairs.

### Assessment of Publication Bias

Possible publication bias was investigated using funnel plots and Egger’s test in cases of ten or more studies, as per the protocol.

### Visualisation

All data-analysis was conducted in R version 4.3.1 using the “meta”, “dmetar”, “metafor”, “tidyverse”, “readxl”, “ggplot2”, “ggeasy”, “reshape2”, “DescTools”, “tm”, “stringr”, “esquisse”, “dplyr”, and “flextable” packages.

## Results

### Study Selection

Initial searches on 11^th^ October 2022 identified 8,318 unique publications across 8 databases. From this, 141 manuscripts underwent full-text screening (Figure 1). A further 115 manuscripts were excluded, leaving 26 papers for full data extraction. The update search (23^rd^ October 2023) identified 1,005 unique publications, leaving 137 for full-text screening and subsequently 5 manuscripts for full data extraction. Across both searches a total of 31 manuscripts met the inclusion criteria.

**Figure 1.**
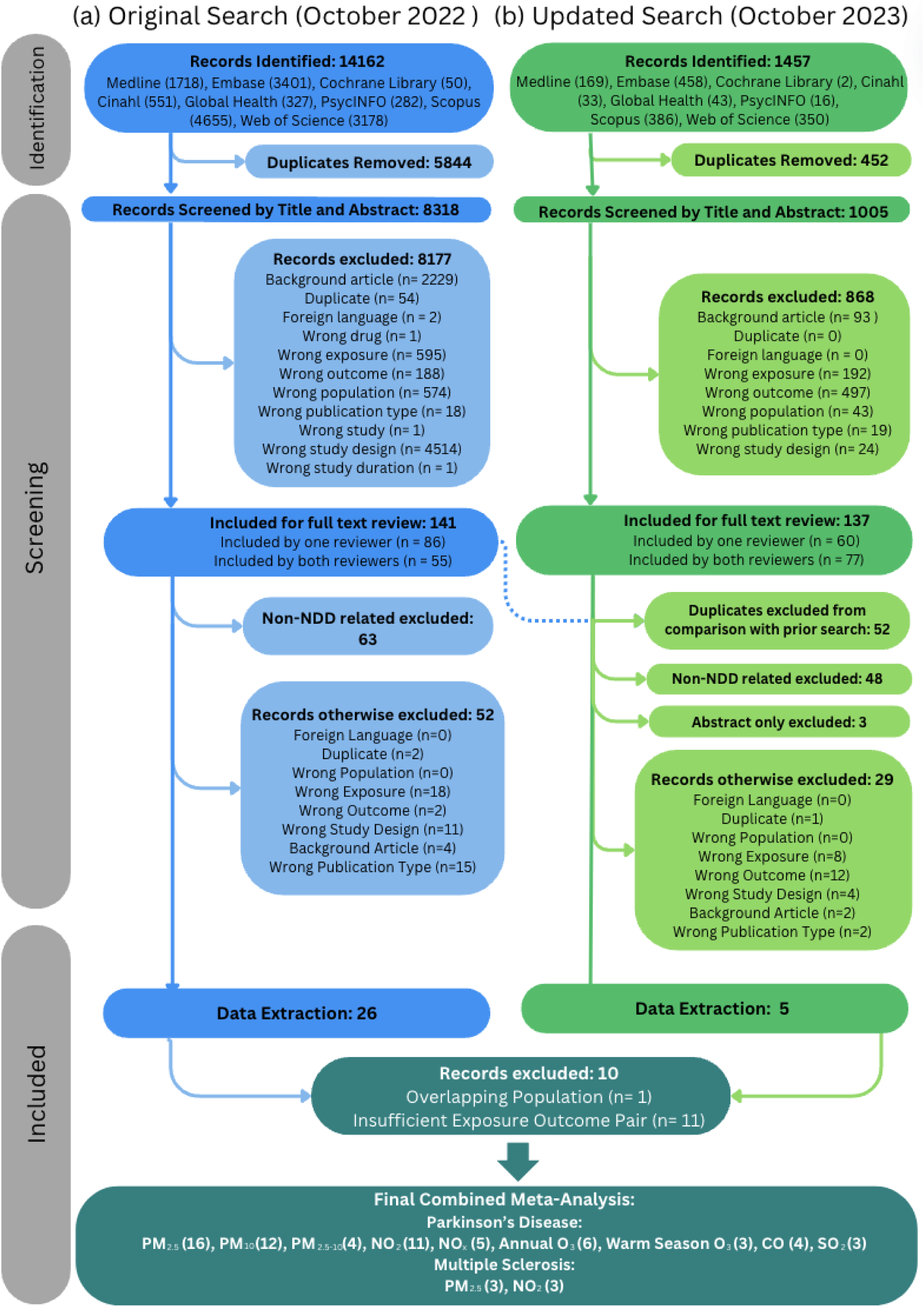
PRISMA Flow Diagram (Exclusions reflect Reviewer 1’s reason where consensus existed but rationales differed)

### Study Characteristics

Of the 31 publications included, 15 pollutants were examined; 21 reported on PM_2.5_, 17 on PM_10_, 16 on NO_2_, 10 on NO_x_ and 7 each for CO, PM_2.5-10_ and annual O_3_. PD, MS and MND incidence was reported as outcomes in 22, 5 and 6 papers, respectively, with some studies including multiple outcomes (Table 3). All studies that investigated incidence of MND focused on ALS only and no other MND disease groups. Outcomes were defined by medical record databases (18), hospital reports (11), medical prescriptions (8), insurance databases (5), hospital databases (2), or insurance claims (1) with many studies using multiple sources to define outcomes. Exposure assessment methods varied across studies, with the most popular methods being data from Land Use Regression (11), chemical transport models (7), spatiotemporal models (6), dispersion models (4), fixed site monitoring (3), and hybrid model (2), with some studies using more than one method depending on pollutant. The majority of studies were from North America [US (10), Canada (4)] followed by Europe [Netherlands (2), Denmark (2), Italy (2), Finland (1) and Sweden (1)], South-East Asia [South Korea (2), China (2), Taiwan (2)] and Middle East [Israel (1)]. The ages of participants ranged from 19 to 109 and follow-up periods ranged from 2 to 25 years. Among these studies, three included only women, one included only men and 27 included both men and women. Ethnicity was reported in 11 of 31 papers mostly from the US, and not reported in any studies from Asia and Europe. There were 17 case-control studies, 15 cohort studies, with one study employing a cohort design to examine PD incidence and a case-control design to examine MS incidence.^46^ No cross-sectional or ecological studies met the inclusion criteria.

**Table 3.** All Studies Included in Systematic Review.

| Author/<br>Year | Country | Study<br>Design | Study Name/<br>Population Size<br>(n) | Sample<br>Size | Exposure | Outcome | Outcome<br>Ascertainment<br>Method | Min/Max<br>Age | Study Period<br>(Follow-up) | Exposure<br>Assessment<br>Method | Source | Spatial<br>Resolution | Confounders<br>(Bolded<br>indicates<br>included in<br>Extracted Effect<br>Estimate) |
| --- | --- | --- | --- | --- | --- | --- | --- | --- | --- | --- | --- | --- | --- |
| (Andrew<br>et al.,<br>2022) | USA | Case-Control | SYMPHONY<br>Integrated<br>Dataverse (NA) | 104,796 | Lead | ALS | Insurance<br>databases,<br>medical<br>prescriptions | NA/NA | 2013-2019<br>(7 years) | Chemical<br>Transport<br>Modelling* | U.S.<br>Environmental<br>Protection<br>Agency<br>(EPA)'s<br>National<br>Emissions<br>Inventory<br>(NEI)* | Zip Code | **Age, Sex** |
| (Bai et<br>al., 2018) | Canada | Cohort | Ontario<br>Population<br>Health and<br>Environment<br>Cohort<br>(2,824,478) | 2,824,478 | PM<sub>2.5</sub><br>NO<sub>2</sub><br>Warm<br>Season O<sub>3</sub> | MS | Hospital<br>reports;<br>insurance<br>claims | NA/NA | 2001-2013<br>(12.75 years) | Global<br>Atmospheric<br>Chemistry<br>Transport<br>Model | GEOS-Chem<br>CTM (PM<sub>2.5</sub>)<br>Environment<br>Canada's<br>National Air<br>Pollution<br>Surveillance<br>Network (NO<sub>2</sub>)<br>Canadian and<br>Hemispheric<br>Regional<br>Ozone and NO<sub>x</sub><br>System (Warm<br>Season<br>O<sub>3</sub>) | 1x1km<br>(PM<sub>2.5</sub>)<br>10x10km<br>(NO<sub>2</sub>)*<br>1x1km<br>(Warm<br>Season<br>O<sub>3</sub>)* | **Age, Sex,**<br>**Smoking,**<br>**Education,**<br>**Income,**<br>**Neighborhood**<br>**level recent**<br>**immigrants,**<br>**Unemployment**<br>**rate, Annual**<br>**household**<br>**income,**<br>**urban/rural,**<br>**Comorbidities**<br>**(diabetes,**<br>**hypertension,**<br>**traumatic brain**<br>**injury,**<br>**infectious**<br>**mononucleosis),**<br>**Latitude** |
| (Cerza et<br>al., 2018) | Italy | Cohort | Rome<br>Longitudinal<br>Study<br>(1,037,895) | 1,008,253 | PM<sub>2.5</sub><br>PM<sub>10</sub><br>PM<sub>2.5-10</sub><br>NO<sub>2</sub> | PD | Medical<br>prescriptions;<br>hospital<br>records | NA/NA | 2008-2013<br>(6 years) | Land Use<br>Regression<br>Models | ESCAPE | 100x100m* | **Age, Sex,**<br>**Education,**<br>**Socioeconomic**<br>**position,** |
|  |  |  |  |  | NO<sub>x</sub><br>Annual O<sub>3</sub> |  |  |  |  |  |  |  | **Marital status,**<br>**Place of birth,**<br>**Comorbidities**<br>**(cardiovascular**<br>**disease)** |
| (C.-Y.<br>Chen et<br>al., 2017) | Taiwan | Case-Control | Taiwan National<br>Health Insurance<br>Program<br>(n=1,000,000) | 5,300 | PM<sub>10</sub><br>NO<sub>2</sub><br>NO<sub>x</sub><br>Annual O<sub>3</sub><br>CO<br>SO<sub>2</sub> | PD | Hospital<br>reports | NA/NA | 2000-2013<br>(14 years) | Taiwan<br>EPA<br>monitoring<br>stations | Environmental<br>Protection<br>Administration<br>(EPA) of<br>Taiwan | 76<br>monitoring<br>stations<br>across<br>Taiwan* | **Age, Sex,**<br>**urban/rural,**<br>**comorbidities**<br>**(diabetes,**<br>**hypertension,**<br>**stroke,**<br>**depression,**<br>**renal disease,**<br>**dementia, head**<br>**injury, alcohol-**<br>**related disease;**<br>**tobacco use**<br>**disorder; sleep**<br>**disorder)** |
| (H. Chen<br>et al.,<br>2017) | Canada | Cohort | Ontario's<br>Registered<br>Persons<br>Database<br>(2,165,268 PD;<br>4,372,720 MS) | 2,165,268<br>(PD)<br>4,372,720<br>(MS) | PM<sub>2.5</sub><br>NO<sub>2</sub> | PD<br>MS | Hospital<br>reports;<br>physician<br>reports;<br>medical<br>prescription | 55/85 for<br>PD;<br>20/50 for<br>MS | 2001-2012<br>(12 years) | Global<br>Atmospheri<br>c Transport<br>Model<br>(PM<sub>2.5</sub>).<br>Land Use<br>Regression<br>Model<br>(NO<sub>2</sub>) | Global<br>Atmospheric<br>Chemistry<br>Transport<br>Model (PM<sub>2.5</sub>)<br>Environment<br>Canada's<br>National Air<br>Pollution<br>Surveillance<br>Network (NO<sub>2</sub>) | 1x1km<br>(PM<sub>2.5</sub>)<br>30x30m<br>(NO<sub>2</sub>)* | **Age, Sex,**<br>**Education,**<br>**Income,**<br>**Unemployment**<br>**rate, % recent**<br>**immigrants,**<br>**urban/rural,**<br>**Residential**<br>**proximity to**<br>**major**<br>**roadways,**<br>**Comorbidities**<br>**(diabetes,**<br>**hypertension,**<br>**coronary heart**<br>**disease, stroke,**<br>**congestive heart**<br>**failure,**<br>**arrhythmia,**<br>**traumatic brain**<br>**injury)** |
| (Filippini | Italy | Case- | ALS Center of | 132 | PM<sub>10</sub> | ALS | Hospital | NA/NA | 1994-2015 | CALINE4 | Local | ~ 100x | **Age, Sex** |
| et al., 2021) |  | Control | Modena University Hospital (145) |  |  |  | reports |  | (NA) | Air Quality Dispersion Model | Vehicular Traffic Reports | 100m* |  |
| (Hedström et al., 2023) | Sweden | Case-Control | Epidemiological Investigation of Multiple Sclerosis (EIMS) and Genes and Environment in Multiple Sclerosis (GEMS) (n=125,268) | 125,268 | NO<sub>x</sub> | MS | Hospital database | NA/NA | 2005-2018 (14 years) | Chemical Transport Modeling | Nordic WelfAir project | 100x100m for road traffic, 200x200m for RWC, shipping and large point sources, 500x500m for other sources | **Age, Sex, BMI, Smoking, Education, Income, Occupational Socioeconomic status, Ancestry,** |
| (Karakis et al., 2023) | Israel | Case-Control | Clalit Health Services (CHS) HMO (n=34,667) | 34,667 | PM<sub>2.5</sub><br>PM<sub>10</sub><br>NO<sub>2</sub><br>Annual O<sub>3</sub><br>SO<sub>2</sub> | PD | Clinician reports; medical prescription | 21/109 | 2000-2020 (8.7 years cases; 8.9 years for controls) | Proximal Monitors to residential address | Monitoring Stations Managed by the Ministry of Environmental Protection | 20km | **Age, Sex, Immigration status; Education, Income, Ethnicity, Neighbourhood level, Proportion of years in follow-up with monitored pollutants' levels out of total years in follow-up, Number of monitoring stations within a 20 km radius around the subject's residence** |
| (Kirrane et al., 2015) | USA | Cohort | Agricultural Health Study (84,739) | Iowa (53,219)<br>North Carolina (29,716) | PM<sub>2.5</sub><br>Annual O<sub>3</sub><br>Warm Season O<sub>3</sub> | PD | Clinician report | NA/92 | 1993-2010 (17 years) | Hierarchical bayesian model combining EPA Air Quality System and Community Multiscale Air Quality model | U.S. Environmental Protection Agency's (EPA) Air Quality System (AQS)* | 12x12km | **Age, Sex, Smoking, Pesticide** |
| (Kwon et al., 2024) | USA | Case-Control | Parkinson's Environmental and Genes (PEG) Study (2,899) | 1,671 | PM<sub>2.5</sub><br>CO | PD | Hospital reports; clinician reports | NA/NA | 2001-2017 (17 years) | Chemical Transport Model (PM<sub>2.5</sub>) Gaussian plume dispersion model (CO) | MERRA-2 meteorological data from NASA's Global Modeling and Assimilation Office (PM<sub>2.5</sub>)<br>1 Camino Real Study (CO) | 1km (PM<sub>2.5</sub>)<br>1500m (CO) | **Age, Sex, Race, Education, Smoking, Pesticide, Study wave** |
| (P.-C. Lee, Liu, et al., 2016) | Taiwan | Case-Control | Taiwan National Health Insurance Program (23,000,000) | 55,585 | PM<sub>10</sub><br>NO<sub>x</sub><br>Annual O<sub>3</sub><br>CO | PD | Hospital reports; medical prescription | NA/NA | 2007-2009 (2 years) | Bayesian Maximum Entropy models |  | 5x5km (counties/cities with less dense air monitoring networks<br>1x1km for counties/cities with denser air monitoring networks | **Age, Sex, insurance premium, urban/rural, comorbidities (Charlson Comorbidity Index), year of diagnosis** |
| (P.-C. Lee, Raaschou-Nielsen, et al., 2016) | Denmark | Case-Control | PASIDA (Parkinson's in Denmark) Study (7,134) | 3,013 | NO<sub>2</sub><br>NO<sub>x</sub><br>CO | PD | Medical Records | NA/NA | 2008-2010 (3 years) | Danish Air GIS Modeling System | Taiwanese Environmental Protection Agency | 5km x 5km for counties/cities with less dense air monitoring networks and a size of 1 km × 1 km for counties/cities with denser air monitoring networks | **Age, Sex, Family history of PD, Smoking, Education, occupational history; year of birth** |
| (H. Lee et al., 2022) | South Korea | Cohort | National Health Insurance Service-National Sample Cohort (1,000,000) | 313,355 | PM<sub>2.5</sub><br>PM<sub>10</sub> | PD | Hospital reports; medical prescription | 19/87 | 2007-2015 (9 years) | Pointwise national-scale model estimated from 300 geographic variables and a spatial correlation structure based on regulatory monitoring data | Seoul Public Health and Environment Research Institute* | district level (~ 394.2x 394.2km)* | **Age, Sex, BMI, smoking, alcohol, activity, education, income, gross regional domestic product in 2005, frequency of healthcare utilization, insurance type, urban rural, Comorbidities (fasting glucose, total cholesterol, traumatic brain injury and mental disorders, Charlson Comorbidity Index), residential province, medication history, percentage over 65** |
| (Liu et al., 2016) | USA | Case-Control | National Institutes of Health - American Association of Retired Persons Diet and Health | 4,869 | PM<sub>2.5</sub><br>PM<sub>10</sub><br>NO<sub>2</sub> | PD | Physician report | 50/71 | 1995-2006 (11 years) | Regionalized national universal kriging framework (PM<sub>2.5</sub>, PM<sub>10</sub>); | U.S. Environmental Protection Agency's (EPA) Air Quality System (AQS)* | 25x25km (PM<sub>2.5</sub>)*<br>25x25km (PM<sub>10</sub>)*<br>30x30m (NO<sub>2</sub>)* | **Age, sex, smoking, caffeine, activity, education, race, region of US.** |
|  |  |  | Study (566,398) |  |  |  |  |  |  | land-use regression model (NO<sub>2</sub>) |  |  |  |
| (Malek et al., 2015) | USA | Case-Control | West Pennsylvania Counties (144) | 132 | Metals<br>Aromatic Solvents<br>Organic/Chlorinated Solvents<br>Pesticides<br>Other Hazardous Air Pollutants | ALS | Clinical History | NA/NA | 2008-2011 (3 years) | US EPA National Scale Air Toxics Assessment data | U.S. Environmental Protection Agency (EPA) National-Scale Air Toxics Assessment | Census Tract Level* | **Age, sex, smoking, family history of ALS, education, race.** |
| (Palacios, Fitzgerald, Hart, et al., 2014) | USA | Cohort | Nurses' Health Study (121,700) | 111,769 | PM<sub>2.5</sub><br>PM<sub>10</sub><br>PM<sub>2.5-10</sub> | PD | Medical Records | NA/NA | 1990-2008 (19 years) | Spatial Temporal Models | US EPA National Air Toxics Assessments (NATA) | Census Tract Level* | **Age, sex, smoking, caffeine, tract level income, housing value, urban rural, geographical region, ibuprofen use** |
| (Palacios, Fitzgerald, Roberts, et al., 2014) | USA | Cohort | Nurses' Health Study (121,700) | 97,430 | Antimony<br>Arsenic<br>Cadmium<br>Chromium<br>Lead<br>Manganese<br>Mercury<br>Nickel | PD | Medical Records | NA/NA | 1990-2008 (18 years) | Complex Dispersion Model used by National Air Toxics Assessments (NATA) | EPA's Air Quality System (AQS) | Not Reported | **Age, sex, smoking, urban rural, population density** |
| (Palacios, Fitzgerald, Hart, et al., 2017) | USA | Cohort | Health Professionals Follow-Up Study (51,529) | 50,352 | PM<sub>2.5</sub><br>PM<sub>10</sub><br>PM<sub>2.5-10</sub> | PD | Medical Records | NA/NA | 1988-2010 (22 years) | Spatial Temporal Models | Environmental Protection Agency (EPA)'s Air Quality System (AQS) | Not Reported | **Age, sex, smoking, caffeine, urban rural, time period, geographical** |

|  |  |  |  |  |  |  |  |  |  |  |  |  | region |
| --- | --- | --- | --- | --- | --- | --- | --- | --- | --- | --- | --- | --- | --- |
| (Palacios , Fitzgerald, Munger, et al., 2017) | USA | Cohort | Nurses' Health Study (121,700)<br>Nurses' Health Study II (116,671) | 112,595<br>102,583 | PM<sub>2.5</sub><br>PM<sub>10</sub><br>PM<sub>2.5-10</sub> | MS | Medical Records | NA/NA | 1988-2004 (7 years)<br>1988 - 2007 (20 years) | Spatial Temporal Models | U.S. Environmental Protection Agency (EPA)'s National Emissions Inventory (NEI) | 500x500m* | **Age, sex, BMI, smoking, tract level income, urban rural, calendar year, geographical region, ethnicity, latitude at age 15; UV index** |
| (Parks et al., 2022) | Denmark | Case-Control | Danish National Hospital Registrar (24,066) | 23,232 | PM<sub>2.5</sub><br>NO<sub>x</sub><br>Annual O<sub>3</sub><br>CO<br>Elemental Carbon | ALS | Medical Records | NA/NA | 1989-2013 (25 years) | Spatial Temporal Models | DEHM-UBM-AirGis* | 1x1km* | **Age, sex, education, occupation, marital status, year of birth, vital status, last reported place of residence, place of birth** |
| (Ritz et al., 2016) | Denmark | Case-Control | Danish National Hospital Registrar (6,388) | 3,496 | NO<sub>2</sub><br>NO<sub>x</sub><br>CO | PD | Medical Records | NA/NA | 1996-2009 (13 years) | GIS-based Dispersion Modeling System | AirGIS | 1x1km* | **Age, sex, family history of PD, smoking, education, occupation, year of birth** |
| (Rumrich et al., 2023) | Finland | Case-Control | FINPARK study (170,198) | 160,712 | PM<sub>2.5</sub><br>PM<sub>10</sub> | PD | Insurance database | NA/NA | 1996-2015 (20 years) | Hybrid | Danish Eulerian Hemispheric Model (DEHM) | 1x1km | **Age, sex, occupation, urban rural, occupational exposure to pesticides, comorbidities (asthma/ smoking-related cancer, traumatic head injury), hospitalisation episodes, days** spent in hospital |
| (Seelen et al., 2017) | Netherlands | Case-Control | Prospective ALS in the Netherlands (16,455, 911) | 3,579 | PM<sub>2.5</sub><br>PM<sub>10</sub><br>PM<sub>2.5-10</sub><br>PM<sub>2.5</sub> absorbance<br>NO<sub>2</sub><br>NO<sub>x</sub> | ALS | Medical Records | NA/NA | 2006-2013 (7 years) | Land Use Regression Model | ESCAPE | 100x100m* | Age, sex, BMI, smoking, alcohol, education, income, urban rural. |
| (Shin et al., 2018) | Canada | Cohort | Ontario Population Health and Environment Cohort (2,194,519) | 2,194,519 | PM<sub>2.5</sub><br>NO<sub>2</sub><br>Warm Season O<sub>3</sub> | PD | Medical Prescriptions, Hospital Reports | NA/NA | 2011-2013 (12 years) | Chemical Transport Model | Moderate Resolution Imaging Spectroradiometer of the National Aeronautics and Space Administration's Terra/Aqua satellite (PM<sub>2.5</sub>)<br>Environment Canada's National Air Pollution Surveillance (NAPS) (NO<sub>2</sub>)<br>Fixed site monitors and Canadian Hemispheric and Regional Ozone and NO<sub>x</sub> System (Warm Season O<sub>3</sub>) | 1x1km (PM<sub>2.5</sub>)<br>Within 10km (NO<sub>2</sub>)<br>21x21km (Warm Season O<sub>3</sub>) | Age, sex, neighbourhood level socioeconomic factors (employment, recent immigrant, high school education, income), urban rural, comorbidities |
| (Jo et al., 2021) | South Korea | Cohort | Korean National Health Insurance Service (1,021,208) | 78,830 | PM<sub>2.5</sub><br>PM<sub>10</sub><br>NO<sub>2</sub> | PD | Insurance database, hospital records | NA/NA | 2002-2015 (14 years) | Seoul Research Institute of Public | Seoul Research Institute of Public Health and | District Level | Age, sex, BMI, smoking, alcohol, activity, insurance type, |
|  |  |  |  |  | Annual O<sub>3</sub><br>CO<br>SO<sub>2</sub> |  |  |  |  | Health and Environme<br>nt Hourly<br>Monitoring | Environment |  | comorbidities<br>(hypertension,<br>diabetes,<br>dyslipidemia,<br>traumatic brain<br>injury, kidney<br>disease,<br>congestive heart<br>failure, ischemic<br>heart disease) |
| (Toro et al., 2019) | Netherl<br>ands | Case-Control | Multi-Center<br>Case-Control<br>Study in the<br>Netherland<br>(3003) | 1,290 | PM<sub>2.5</sub><br>PM<sub>10</sub><br>PM<sub>2.5-10</sub><br>NO<sub>2</sub><br>NO<sub>x</sub> | PD | Medical<br>records | NA/NA | 2010-2012<br>(2 years) | Land Use<br>Regression<br>Model | ESCAPE | 100x100m* | **Age, sex, family**<br>**history,**<br>**smoking,**<br>**education,**<br>**neighbourhood**<br>**level income,**<br>**study site** |
| (Willis et al., 2022) | USA | Cohort | Medicare Data<br>(29,000,000) | 5,138,648 | Copper<br>Lead<br>Manganese | PD | Insurance<br>databases,<br>medical<br>records | 70/NA | 1995-2003<br>(9 years) | US EPA<br>Toxic<br>Release<br>Inventory<br>database | Environmental<br>Protection<br>Agency Toxic<br>Release<br>Inventory | County<br>Level | **Age, sex, race** |
| (Z. Yu, Wei, et al., 2021) | China | Cohort | Prospective<br>Cohort Study in<br>Yinzhou District<br>of Ningbo,<br>China (48,908) | 46,839 | PM<sub>2.5</sub><br>PM<sub>10</sub><br>NO<sub>2</sub> | PD | Medical<br>records | NA/NA | 2009-2020<br>(12 years) | Land Use<br>Regression<br>Model | Qingyue<br>environmental<br>protection<br>information<br>technology<br>service | Not<br>Reported | **Age, sex, BMI,**<br>**smoking,**<br>**caffeine,**<br>**alcohol, activity,**<br>**education,**<br>**income, marital**<br>**status** |
| (Yuchi et al., 2020) | Canada | Cohort<br>(PD)<br>Case-Control<br>(MS) | Population Data<br>British<br>Columbia<br>(641,664) | 634,432<br>(PD)<br>7,232<br>(MS) | PM<sub>2.5</sub><br>NO<sub>2</sub> | PD<br>MS | Insurance<br>databases,<br>medical<br>prescriptions,<br>hospital<br>records | 45/83<br>(PD)<br>NA/NA<br>(MS) | 1999-2003<br>(5 years) | Land Use<br>Regression<br>Model | Monitoring<br>Sites | Not<br>Reported | **PD: Age, sex,**<br>**ethnicity,**<br>**education,**<br>**income,**<br>**comorbidities**<br>**(traumatic**<br>**brain injury,**<br>**diabetes,**<br>**hypertension,**<br>**stroke,**<br>**coronary heart** disease, congestive heart failure, arrhythmia)<br>MS: Education, income, ethnicity, comorbidities (traumatic brain injury, diabetes, hypertension, stroke, coronary heart disease, congestive heart failure, arrhythmia) |
| (Z. Yu, Peters, et al., 2021) | Netherlands | Case-Control | Prospective ALS in the Netherlands Study (16,455, 911) | 5,660 | PM<sub>2.5</sub><br>PM<sub>10</sub><br>PM<sub>2.5-10</sub><br>PM<sub>2.5</sub> absorbance<br>NO<sub>2</sub><br>NO<sub>x</sub> | ALS | Medical records | NA/NA | 2006-2013 (9 years) | Land Use Regression Model | Monitoring Sites | 25 sampling sites across 2200km<sup>2</sup> | Age, sex, BMI, smoking, alcohol, education, socioeconomic factors. |
| (Zhu et al., 2023) | China | Cohort | Yinzhou District Cohort (47,516) | 29,025 | PM<sub>2.5</sub><br>PM<sub>10</sub><br>NO<sub>2</sub> | PD | Hospital reports and databases | NA/NA | 2015-2018 (median 5.82 years) | Land Use Regression Model | China's National Environmental Monitoring Center via Qingyue Open Environmental Data Center | 10x10km* | Age, sex, BMI, smoking, diet, alcohol, education, occupation, income, tea consumption |

\*indicates that information was found in a source cited from published study

### Meta-analysis of PD incidence

Of the 22 papers related to PD incidence, four papers^47–50^ were not included in meta-analyses. One was excluded in meta-analyses^48^ because the population studied overlapped with another included paper.^51^ Two others^49,50^ examined only metal exposures, e.g., lead, manganese, copper and there were an insufficient number of studies for these metal exposures to meet the criteria for at least three exposure-outcome pairs. The fourth paper^47^ examined exposure-outcome pairs using categorical and not continuous estimates. That study found that NO_2_ exposure was associated with an increased risk of PD (hazard ratio for highest versus lowest quartile, 1.41; 95% CI, 1.02-1.95), and exposure to PM_2.5_, PM_10_, O_3_, SO_2_, or CO were not statistically significantly associated.

We meta-analysed nine pollutants (PM_2.5_, PM_10_, PM_2.5-10_, NO_2_, NO_x_, annual O_3_, warm season O_3_, CO, and SO_2_) pooling adjusted effect estimates and two pollutants (PM_2.5_ and PM_10_) pooling unadjusted effect estimates (Table 4). For pooled adjusted effect estimates, one paper reported two independent effect estimates (one for the state of Iowa and one for the state of North Carolina in the US), which were included separately.^52^ Meta-analysis of the adjusted effect estimates showed a higher risk (1.06; 95% CI: 1.00-1.12) of PD for every 5 μg/m^3^ increase in long-term exposure in PM_2.5_ (Figure 2A) but this was not true for all study settings (PI 0.95-1.19). There was no evidence of an association with PM_10_ (1.00, 95% CI: 0.93-1.07 per 15 μg/m^3^ increase), PM_2.5-10_ (0.99, 95% CI: 0.97-1.01 per 3 μg/m^3^ increase), NO_2_ (1.01, 95% CI: 0.97-1.05 per 10 μg/m^3^ increase), NO_x_ (1.01, 95% CI: 0.94, 1.08 per 17 μg/m^3^ increase), and annual O_3_ (1.01, 95% CI: 0.94, 1.09 per 12 μg/m^3^ increase) (Figure 2B- 2F). PD risk was not statistically significantly associated with CO exposure (1.03, 95% CI: 0.62, 1.71 per 0.02 mg/ increase), warm season O_3_ (1.09, 95% CI: 0.39, 3.09 per 12 μg/m^3^ increase), and SO_2_ (1.09, 95% CI: 0.78, 1.53 per 3 μg/m^3^ increase); but only three studies were included for each of these three pollutants (Figure 2G-2I).

**Figure 2.**
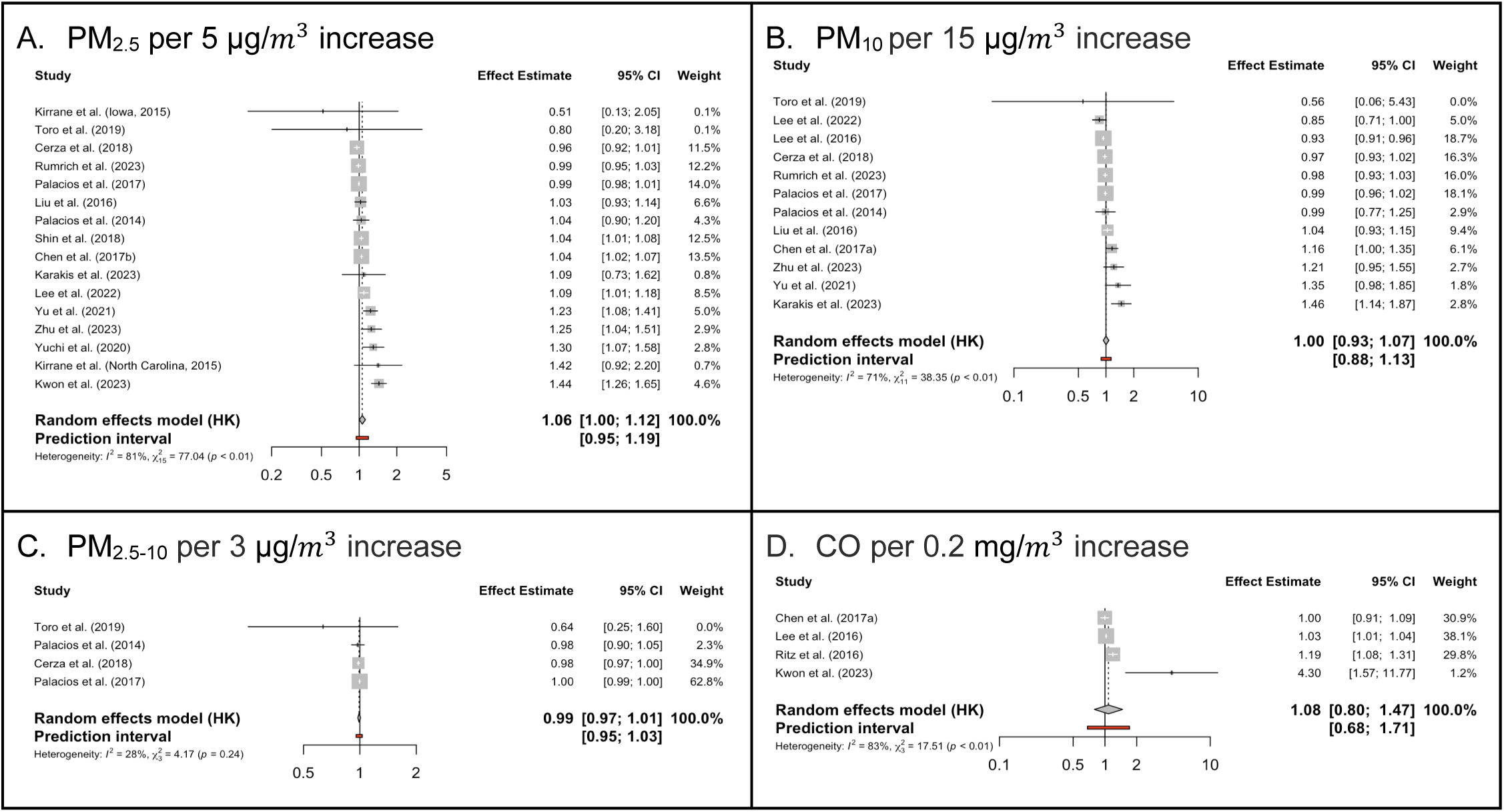
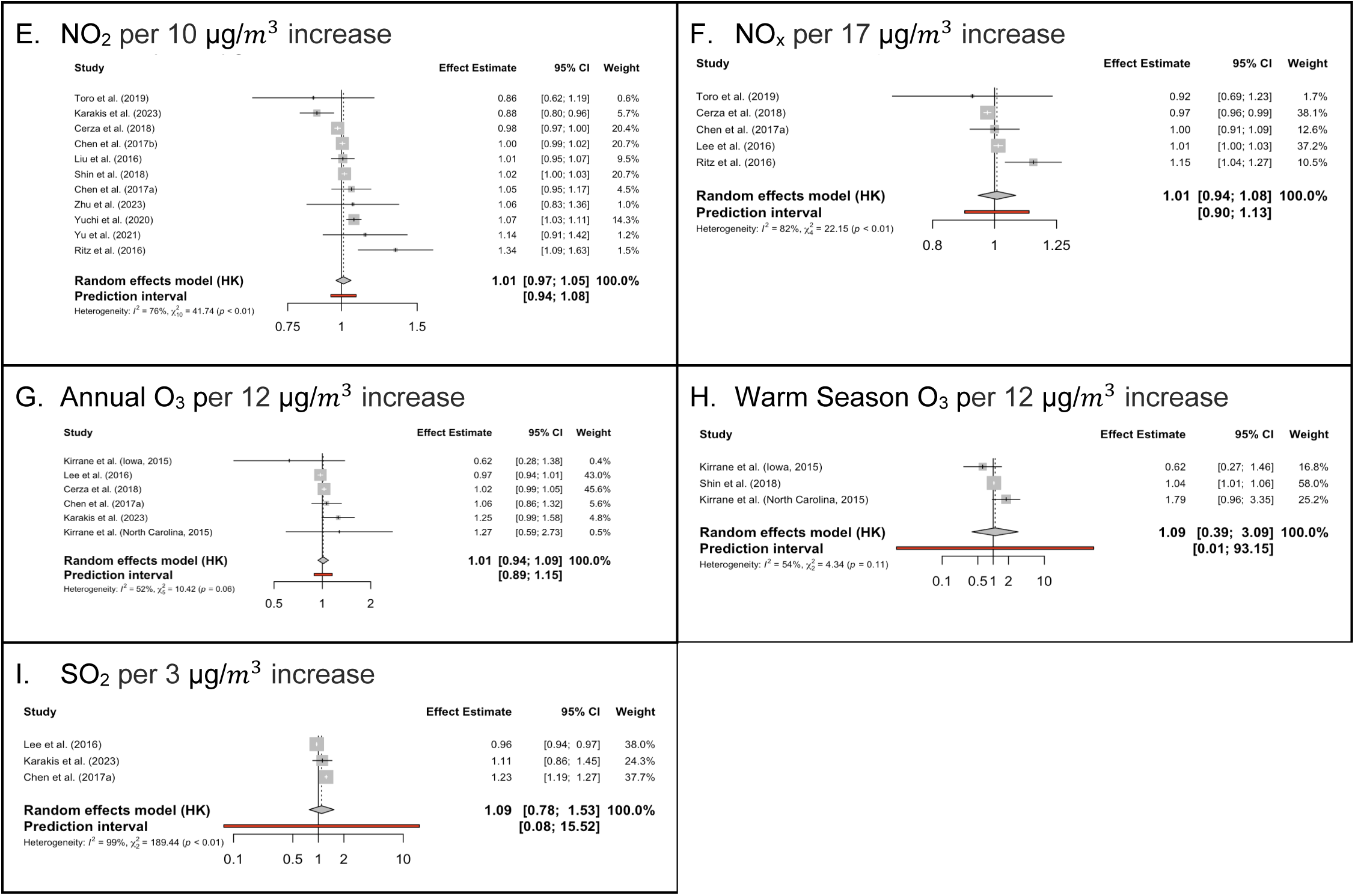
(A – I). Forest Plots (adjusted effect estimates) of the association between ambient air pollutant and Parkinson’s disease incidence. Two studies were published by different first authors with the same surname and are differentiated by using ‘a’ and ‘b’ after the year of publication (C.-Y. Chen et al., 2017a) (H. Chen et al., 2017b).

**Table 4.** Overall adjusted and unadjusted pooled effect estimates per continuous exposure by pollutant, outcome, and number of effect estimates for PD incidence.

| Parkinson's Disease (adjusted effect estimates) |  |  |  |  |
| --- | --- | --- | --- | --- |
| Pollutant | # of Effect Estimates | Overall Adjusted Pooled Effect Estimates | $I^2$ , p-value* | Prediction Interval |
| PM<sub>2.5</sub> | 16 | 1.06 (1.00, 1.12) per 5 $\mu\text{g}/\text{m}^3$ increase | 81%, $\chi^2=77.04$, $p < 0.01$ | 0.95, 1.19 |
| PM<sub>10</sub> | 12 | 1.00 (0.93, 1.07) per 15 $\mu\text{g}/\text{m}^3$ increase | 71%, $\chi^2=38.35$, $p < 0.01$ | 0.88, 1.13 |
| PM<sub>2.5-10</sub> | 4 | 0.99 (0.97, 1.01) per 3 $\mu\text{g}/\text{m}^3$ increase | 28%, $\chi^2= 4.17$, $p = 0.24$ | 0.95, 1.03 |
| NO<sub>2</sub> | 11 | 1.01 (0.97, 1.05) per 10 $\mu\text{g}/\text{m}^3$ increase | 76%, $\chi^2= 41.74$, $p < 0.01$ | 0.94, 1.08 |
| NO<sub>x</sub> | 5 | 1.01 (0.94, 1.08) per 17 $\mu\text{g}/\text{m}^3$ increase | 82%, $\chi^2= 22.15$, $p < 0.01$ | 0.90, 1.13 |
| Annual O<sub>3</sub> | 6 | 1.01 (0.94, 1.09) per 12 $\mu\text{g}/\text{m}^3$ increase | 52%, $\chi^2= 10.42$, $p = 0.06$ | 0.89, 1.15 |
| Warm Season O<sub>3</sub> | 3 | 1.09 (0.39, 3.09) per 12 $\mu\text{g}/\text{m}^3$ increase | 54%, $\chi^2= 4.34$, $p = 0.11$ | 0.01, 93.15 |
| CO | 4 | 1.08 (0.80, 1.47) per 0.2 $\text{mg}/\text{m}^3$ increase | 83%, $\chi^2= 17.51$, $p < 0.01$ | 0.68, 1.71 |
| SO<sub>2</sub> | 3 | 1.09 (0.78, 1.53) per 3 $\mu\text{g}/\text{m}^3$ increase | 99%, $\chi^2= 189.44$, $p < 0.01$ | 0.08, 15.52 |
| Parkinson's Disease (unadjusted effect estimates) |  |  |  |  |
| Pollutant | # of Effect Estimates | Overall Unadjusted Pooled Effect Estimates | $I^2$ , p-value* | Prediction Interval |
| PM<sub>2.5</sub> | 4 | 1.04 (0.85, 1.29) per 5 $\mu\text{g}/\text{m}^3$ increase | 84%, $\chi^2= 18.46$, $p < 0.01$ | 0.70, 1.56 |
| PM<sub>10</sub> | 3 | 0.98 (0.95, 1.02) per 15 $\mu\text{g}/\text{m}^3$ increase | 0%, $\chi^2= 0.46$, $p = 0.80$ | 0.78, 1.24 |

\*p-value is from Cochran's Q (chi-squared) test for heterogeneity

In the meta-analysis for unadjusted effect estimates, no association was identified with PM_2.5_ exposure (1.04 (95% CI: 0.85, 1.29) per 5 μg/m^3^ increase) and PM_10_ (0.98 (95% CI: 0.95, 1.02) per 15 μg/m^3^ increase) (Figure 3).

**Figure 3.**
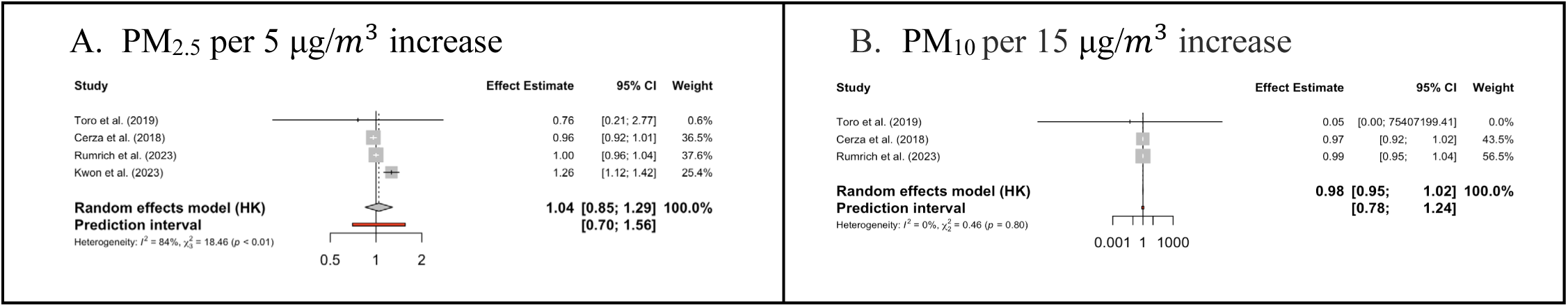
(A-B). Forest Plots (unadjusted effect estimates) of the association between ambient air pollutant and Parkinson’s disease incidence.

**Figure 4.**
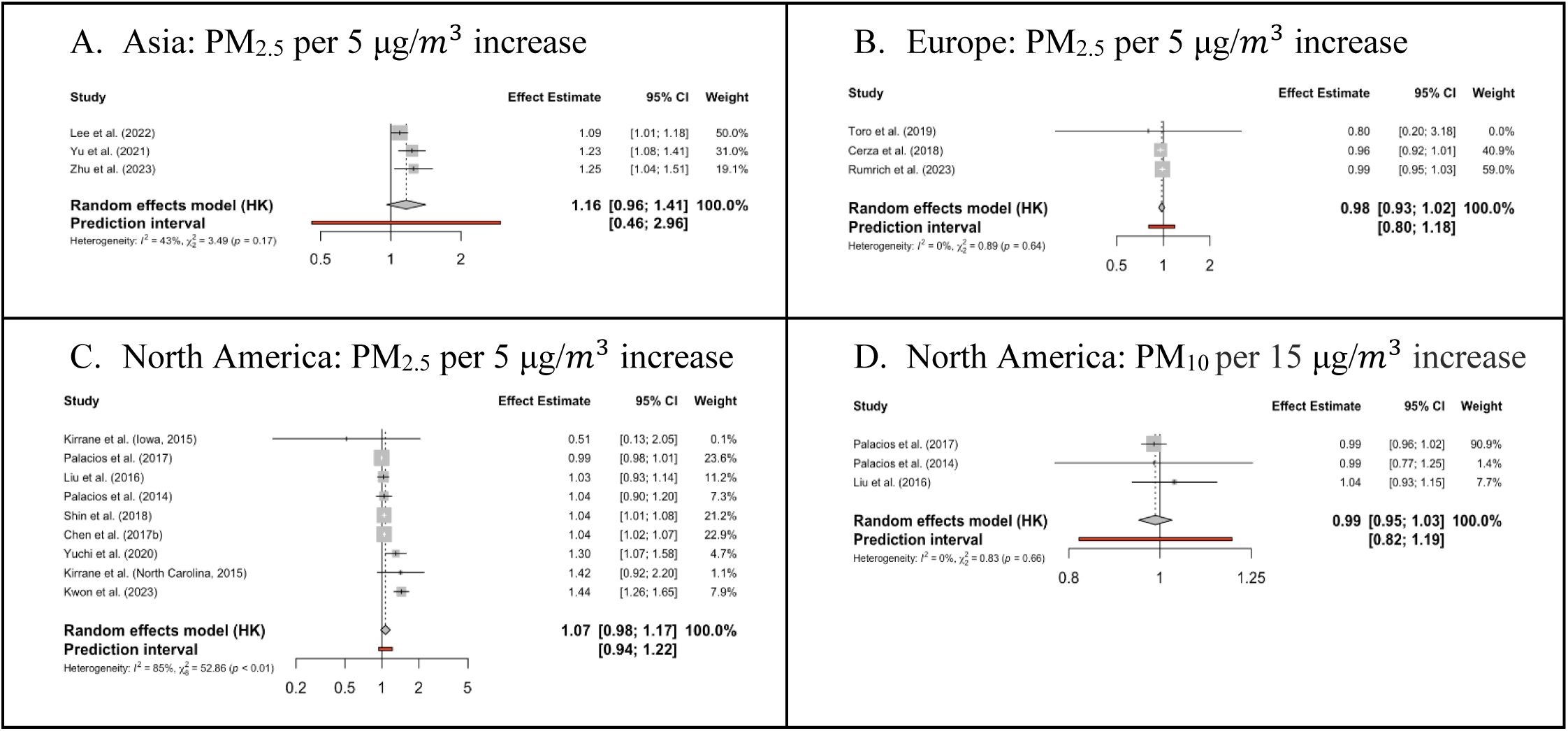
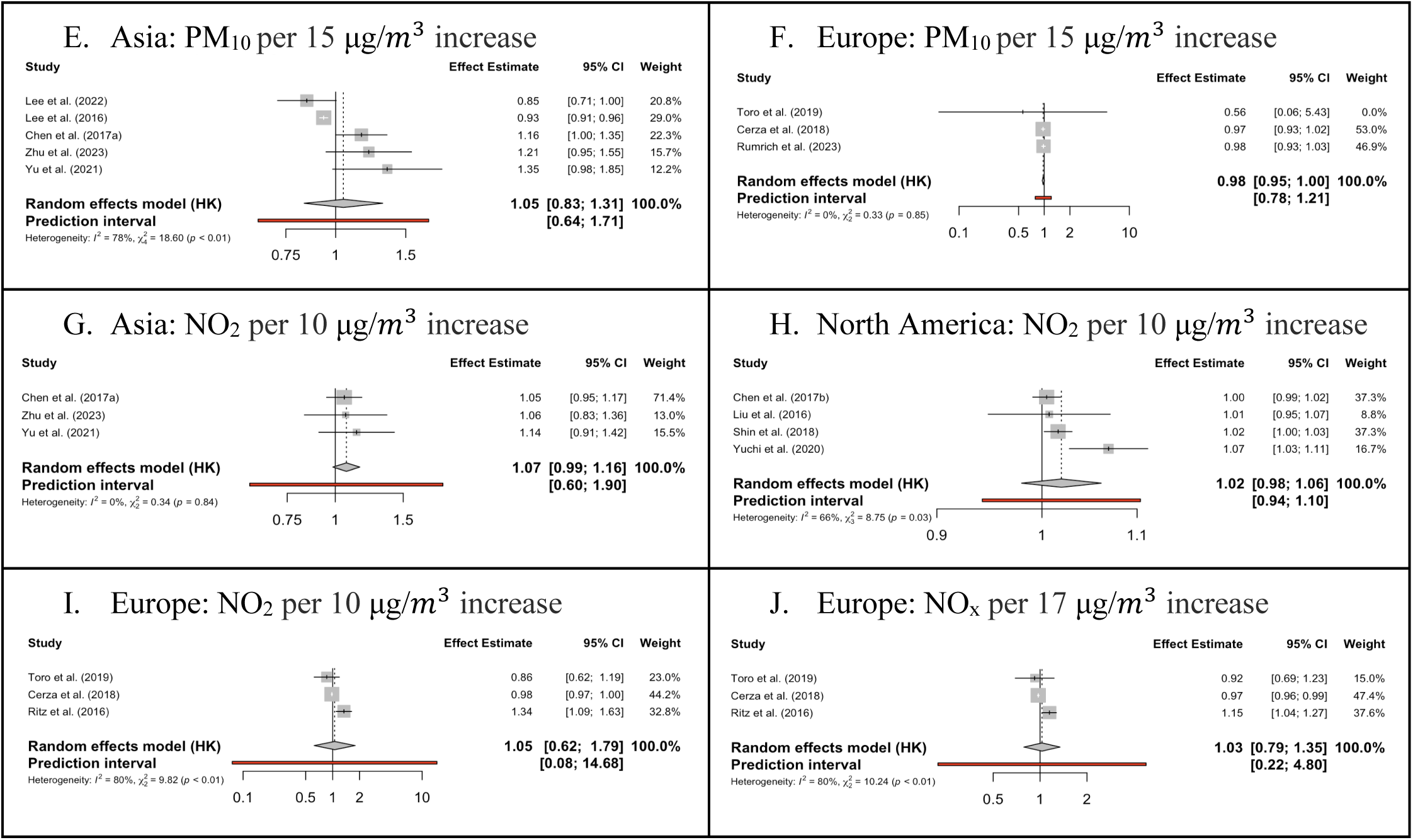
(A-J). Forest Plots (adjusted estimates) of the association between ambient air pollutant and Parkinson’s disease incidence by geographic region.

For adjusted effect estimates, between study heterogeneity was substantial (I^2^ ≥75%) for PM_2.5_ (X^2^ = 77.04, p < 0.01), CO (X^2^ = 17.51, p < 0.01), NO_2_ (X^2^ = 41.74, p < 0.01), NO_x_ (X^2^ = 22.15, p < 0.01) and SO_2_ (X^2^ = 189.44, p < 0.01), moderate (I^2^ = 50-74%) for PM_10_ (X^2^= 38.35, p < 0.01), annual O_3_ (X^2^= 10.42, p = 0.06), and warm season O_3_ (X^2^= 4.34, p = 0.11) and low (I^2^ <50%) for PM_2.5-10_ (X^2^= 4.17, p = 0.24). For unadjusted effect estimates, between study heterogeneity was substantial for PM_2.5_ (I^2^ = 84%, X^2^= 18.46, p < 0.01) and low for PM_10_ (I^2^ = 0%, X^2^= 0.46, p = 0.80).

Where subgroups had three or more effect estimates, meta-analyses were performed. Subgroup analyses by geographical region (North America, Europe and Asia only), study design (cohort study only), exposure assessment methods, and effect estimate type (OR and HR separately) were carried out for adjusted effect estimates for PM_2.5_ and PM_10._ Subgroup analyses were limited to geographic region, study design, and HR effect estimate only for NO_2_, geographic region and OR effect estimate for NO_x_, study design and OR effect estimate only for annual O_3_, and study design for PM_2.5-10_. By geographical region, the largest effect estimates were seen in Asia for PM_2.5_ (1.16, 95% CI: 0.96-1.41), North America for PM_2.5_ (1.07, 95% CI: 0.98-1.17) and Asia for NO_2_ (1.07, 95% CI: 0.99-1.16). None of the subgroups reached statistical significance but the number of studies included in them was rather limited (Figure 5A-E; Table S4 for geographic region; Table S5 for study design and Figure 6; Table S6 for OR and HR effect estimates). For risk of PD from long-term exposure to PM_2.5_ and PM_10_, there was no difference between exposure assessment methods (p = 0.14 and p = 0.12, respectively, Figure S1 and Figure S2).

**Figure 5.**
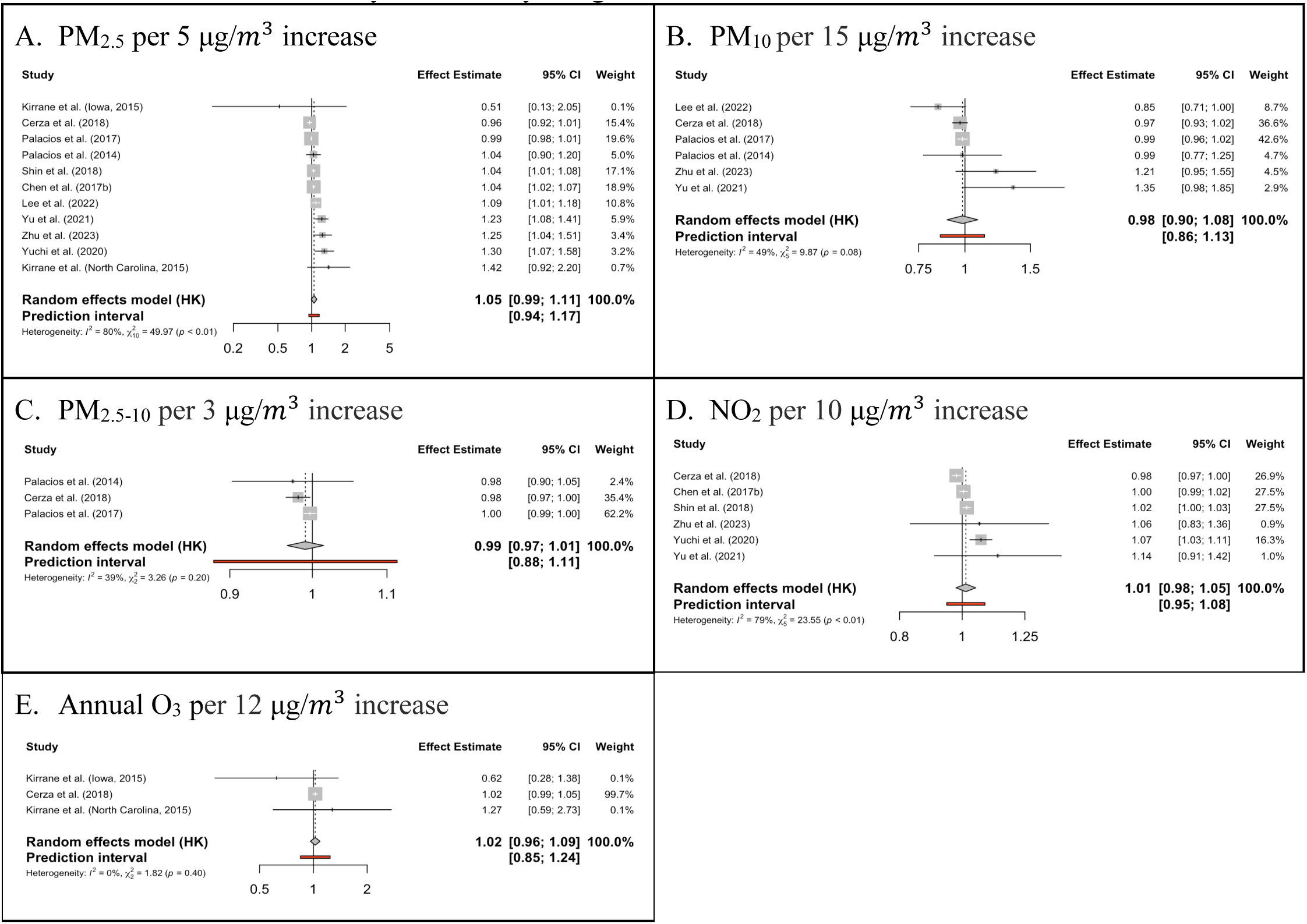
(A-E). Forest Plots (adjusted estimates) of the association between ambient air pollutant and Parkinson’s disease incidence by cohort study design.

**Figure 6.**
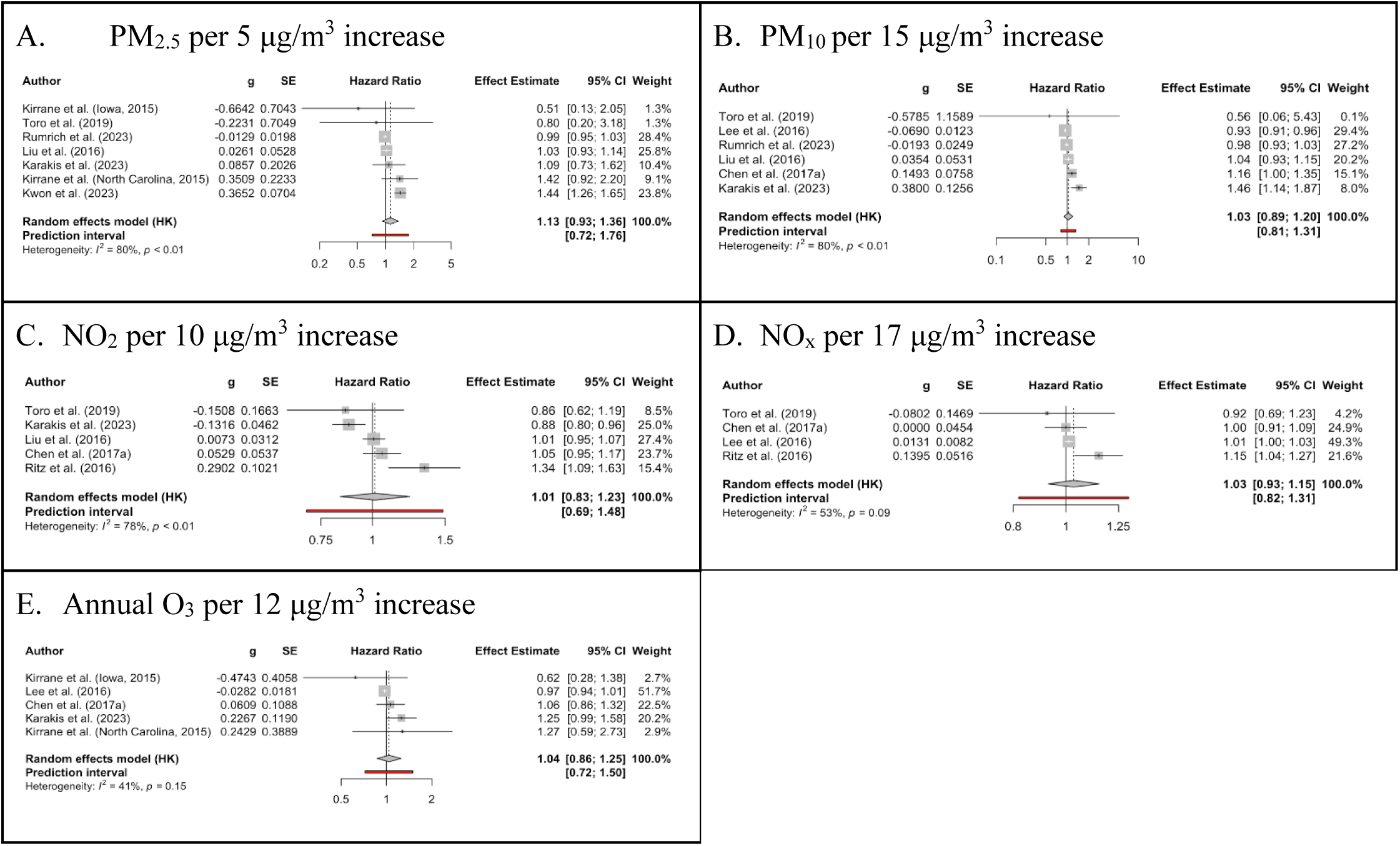
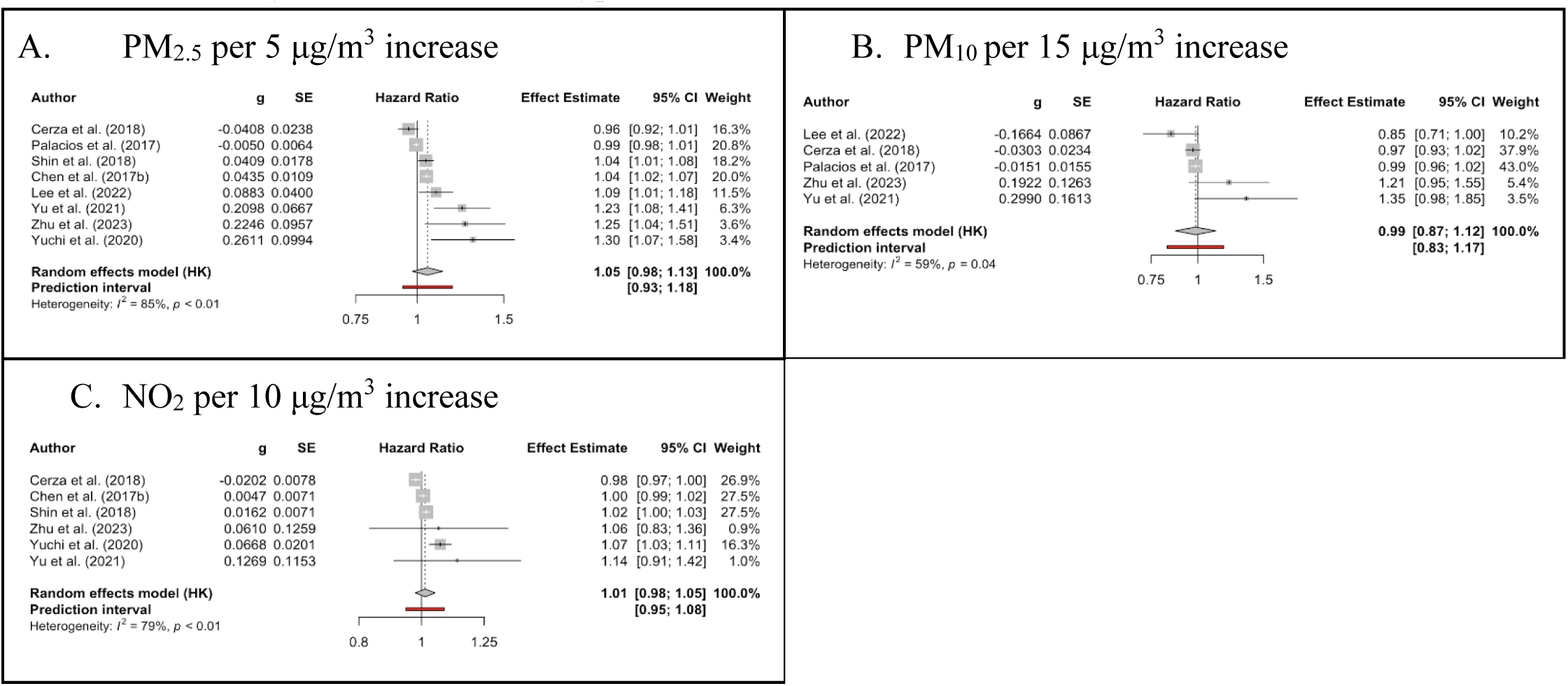
A. Forest Plots (adjusted estimates) of the association between ambient air pollutant and Parkinson’s disease incidence by OR effect estimate type.

Assessment of small study effects was carried out through funnel plots and Egger’s regression for adjusted effect estimates relating to PM_2.5_, PM_10_ and NO_2_ because they had 10 or more studies. There was slight asymmetry in the funnel plots (confirmed by Egger’s) for PM_2.5_ (a = 1.45, p = 0.042) and PM_10_ (a = 1.48, p = 0.048) indicating presence of publication bias (Figure 7 and 8). This was not the case for NO_2_ (a = 0.565, p = 0.54, Figure 9).

**Figure 7.**
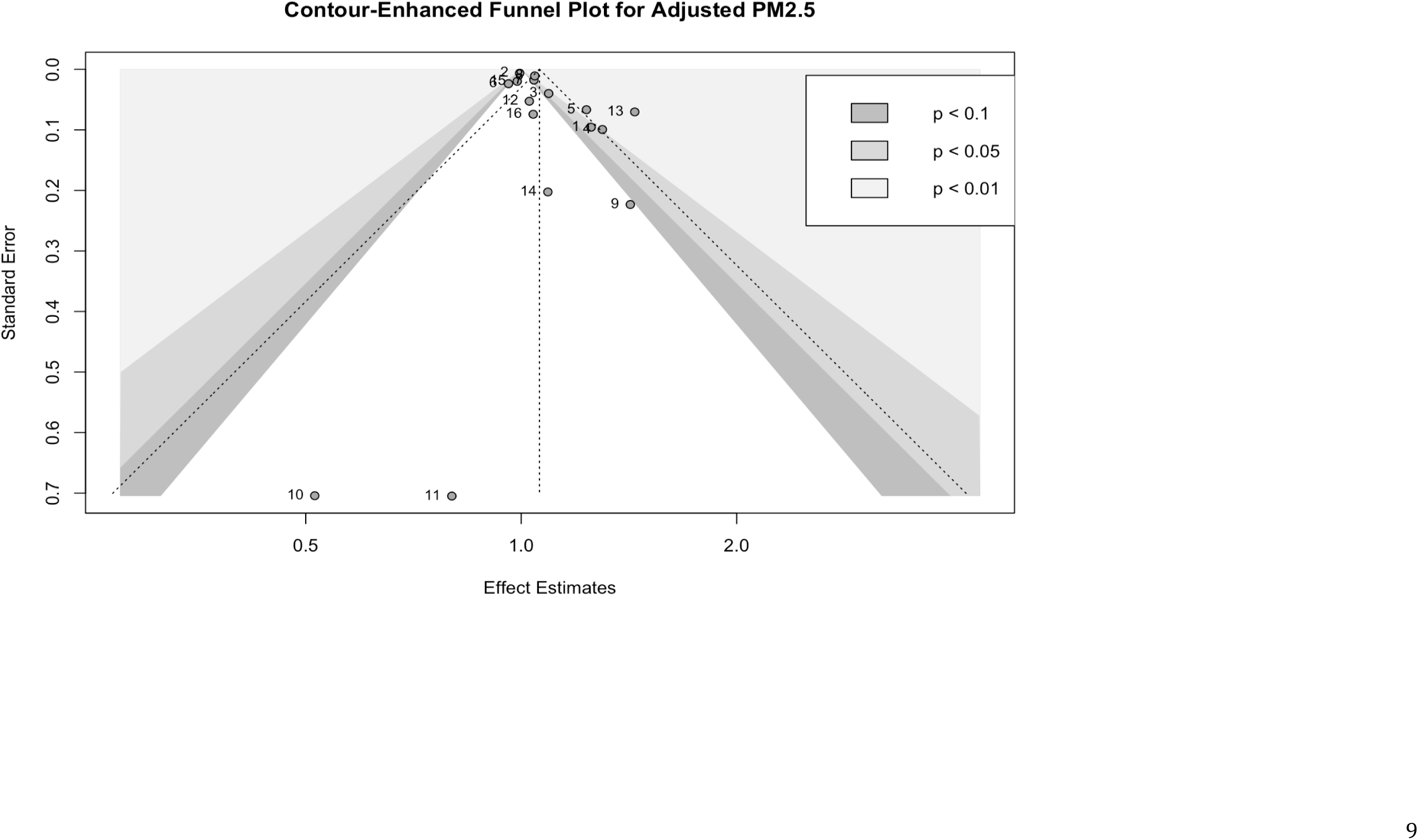
Funnel Plot for Adjusted Continuous PM_2.5_ and Parkinson’s Disease

**Figure 8.**
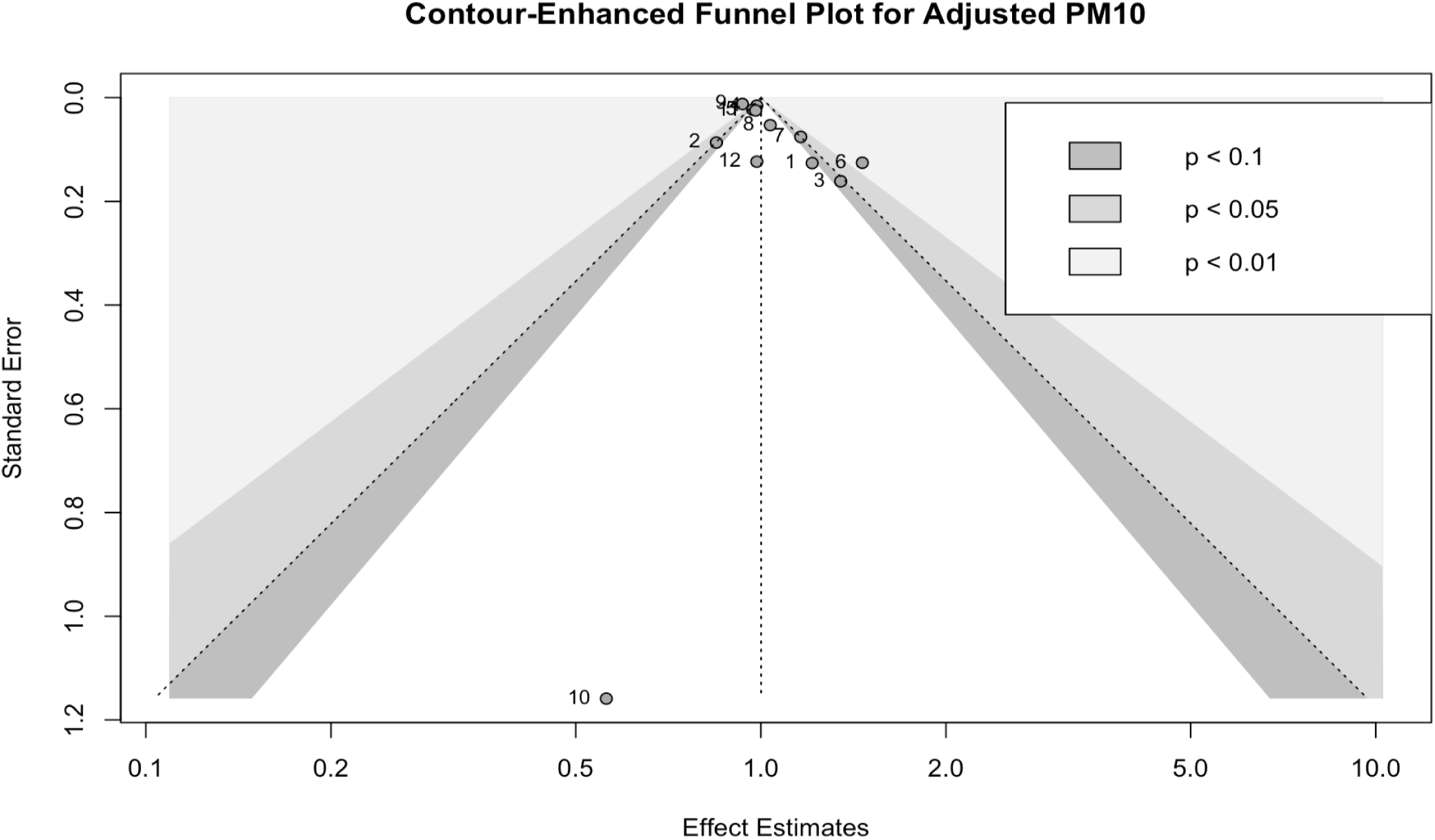
Funnel Plot for Adjusted Continuous PM_10_ and Parkinson’s Disease

**Figure 9.**
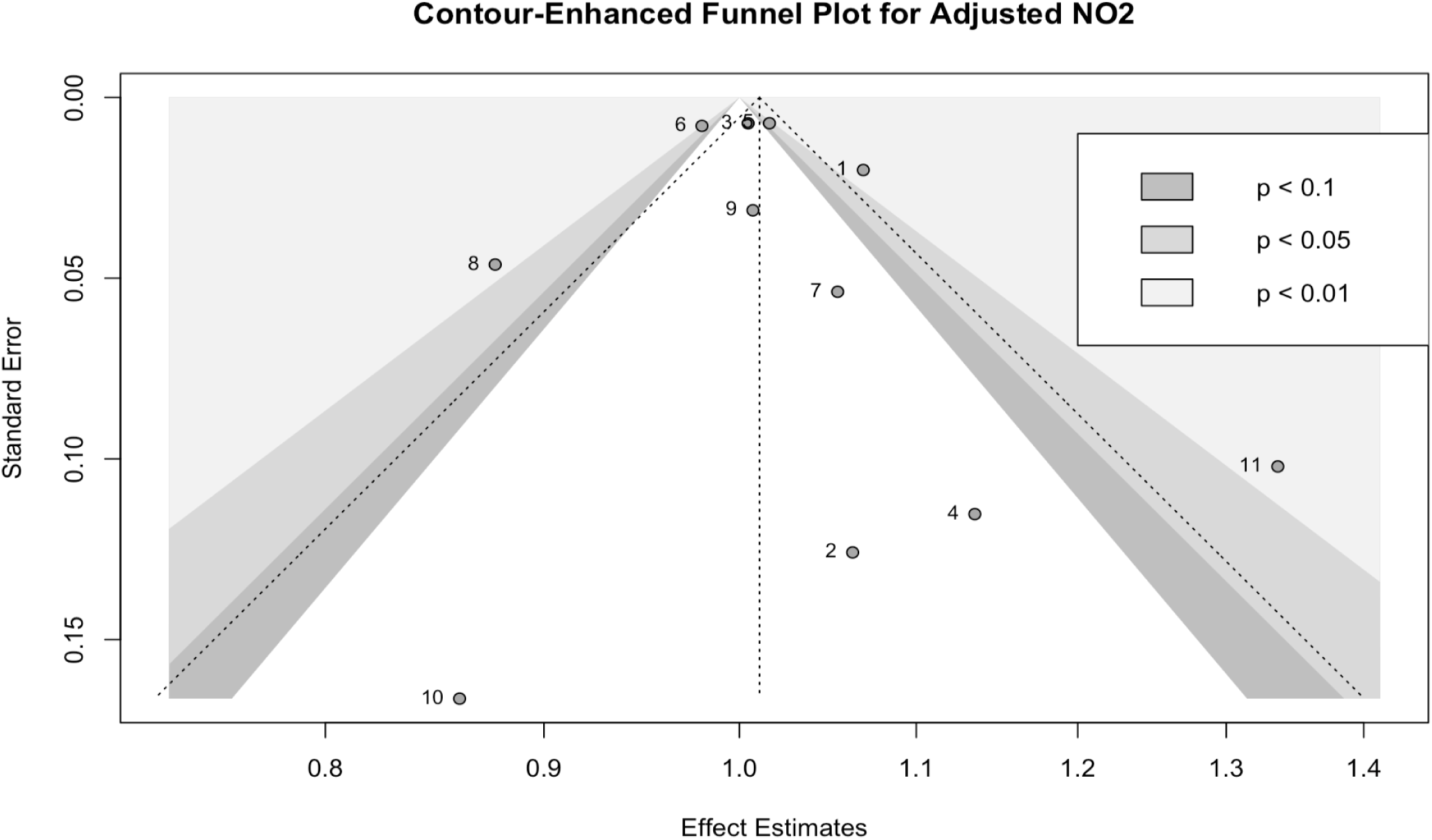
Funnel Plot for Adjusted Continuous NO_2_ and Parkinson’s Disease

Categorical Exposure Response Functions (ERF)

Nine, ten, and six studies reported categorical effect estimates for PM_2.5_, PM_10_, and NO_2_ with PD, though for different ranges of exposures. The visualisations of the ERFs did not show a clear dose-response relationship, with both monotonic and non-monotonic relationships (Figure S3).

### Meta-analysis of MS incidence

Of the five papers (three from Canada^46,53,54^, one from the US^55^, and one from Sweden^56^) examining MS incidence in this systematic review, only the three cohort studies from Canada met inclusion criteria for meta-analysis. Two papers^55,56^ were excluded from the meta-analyses because they examined exposure-outcome pairs using categorical and not continuous estimates. The case-control study from Sweden^56^ found that compared to the lowest quartile, those in the 75-90% and 90% NO_x_ concentration groups had a higher risk of MS (75-90%; 1.06, 95% CI: 1.01-1.11 and 90%; 1.08, 95% CI: 1.03-1.14). By contrast, the

US study ^55^ did not find a consistent association between the lowest and higher quintiles of exposure to PM2.5 (1.04; 95% CI: 0.73-1.50), PM10 (1.11; 95% CI: 0.74-1.87) and PM2.5-10 (1.09; 95% CI: 0.73-1.62) and MS risk.

The three included studies provided a sufficient number of effect estimates for PM_2.5_ and NO_2_ (Table 5). There was no evidence that higher exposure to PM_2.5_ (1.01, 95% CI: 0.77, 1.32 per 5 μg/m^3^ increase) or NO_2_ (0.98, 95% CI: 0.95, 1.01 per 10 μg/m^3^ increase) was associated with higher MS incidence (Figure 10). There was little between study heterogeneity for PM_2.5_ (I^2^ = 34%, X^2^= 3.03, p = 0.22) and NO_2_ (I^2^ = 0%, X^2^ = 0.79, p = 0.67). It was not possible to pool unadjusted effect estimates, carry out subgroup analyses or assess for publication bias due to insufficient number of studies.

**Figure 10.**
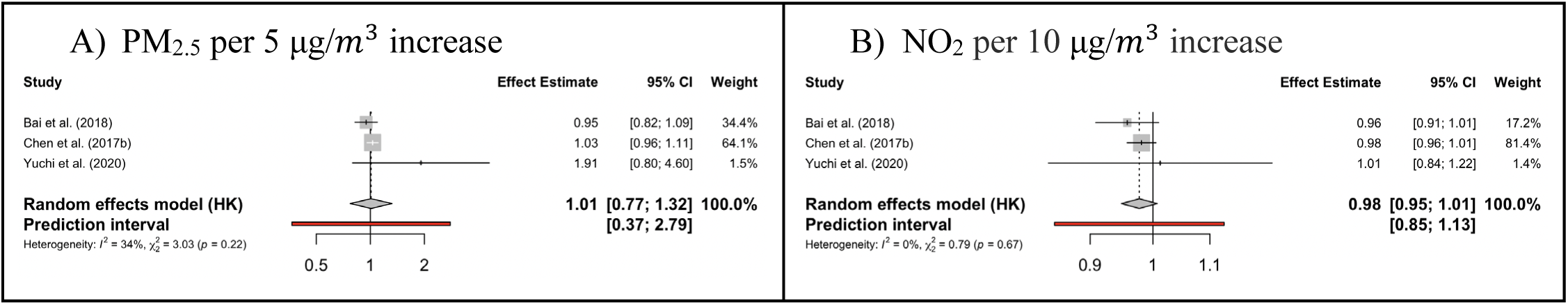
(A-B). Forest Plots (adjusted effect estimates) of the association between ambient air pollutant and Multiple Sclerosis incidence.

**Figure 11.**
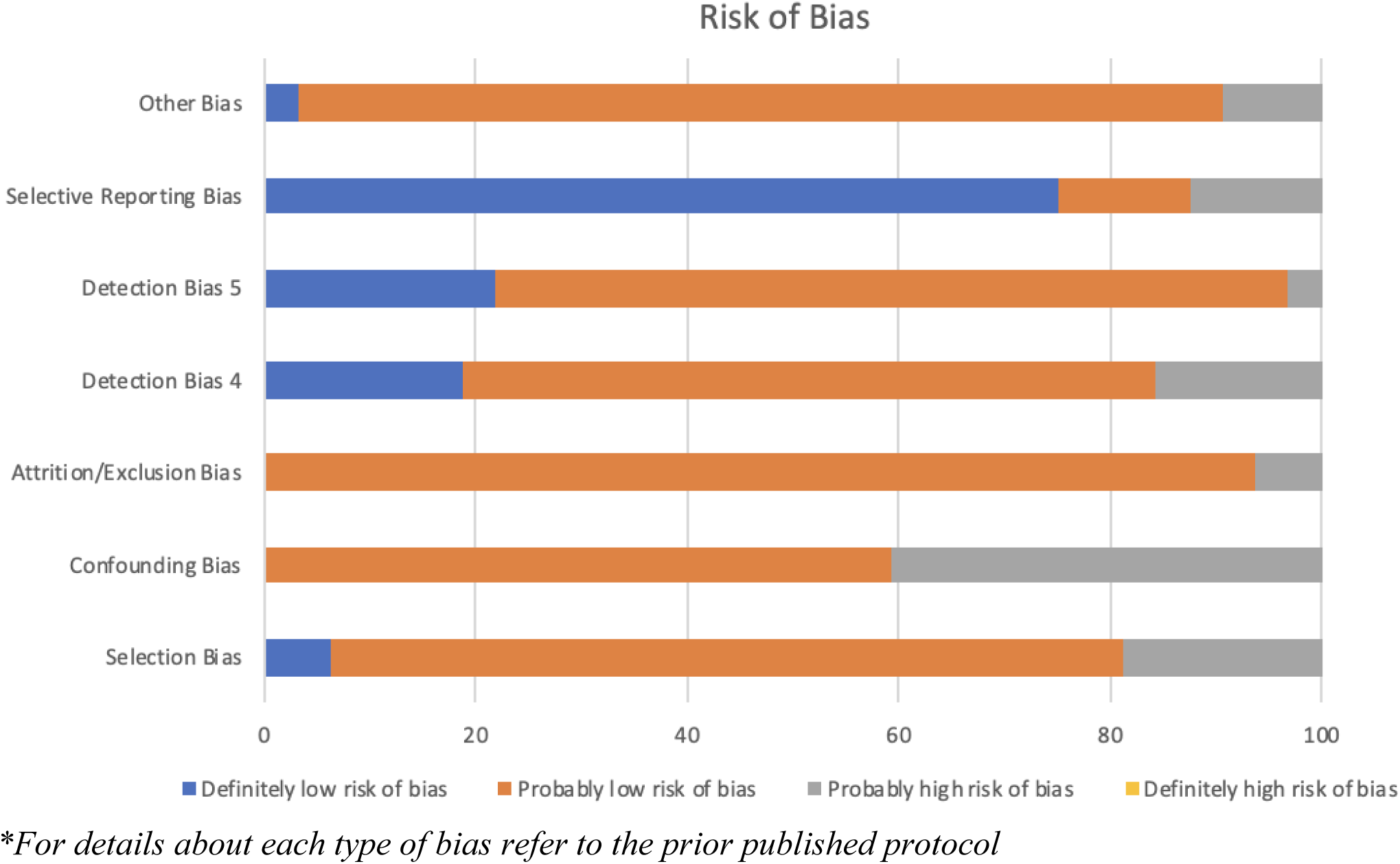
Risk of Bias Results given as a percentage of studies that contained each level of bias* *\*For details about each type of bias refer to the prior published protocol*

**Table 5.** Overall adjusted pooled effect estimates per continuous exposure by pollutant, outcome, and number of effect estimates for MS incidence.

| Multiple Sclerosis |  |  |  |  |
| --- | --- | --- | --- | --- |
| Pollutant | # of Effect Estimates | Overall Adjusted Pooled Effect Estimates | $I^2$, p-value* | Prediction Interval |
| PM<sub>2.5</sub> | 3 | 1.01 (0.77, 1.32) per 5 $\mu\text{g}/\text{m}^3$ increase | 34%<br>($\chi^2 = 3.03$, p=0.22) | 0.37, 2.79 |
| NO<sub>2</sub> | 3 | 0.98 (0.95, 1.01) per 10 $\mu\text{g}/\text{m}^3$ increase | 0%<br>($\chi^2 = 0.79$, p = 0.67) | 0.85, 1.13 |

### Meta-analysis of MND incidence

All six papers ^24,57–61^ included in the systematic review only assessed incidence of ALS, and no other MND diseases. All were case-control studies and exposures did not reach the threshold to conduct a meta-analysis (for continuous exposure) nor to assess the shape of the ERF. The studies were from the US^57,59^, Netherlands^24,61^, Denmark^60^, and Italy.^58^

Among the two studies from the Netherlands, one^24^ found that when compared to Quartile 1 (Q1) of NO_2_ exposure, there was a higher unadjusted and adjusted odds of ALS for concentrations in Q2 (1.33, 95% CI: 1.05-1.61 and 1.38, 95% CI: 1.09-1.76, respectively) and Q4 (1.71, 95% CI: 1.32-2.23 and 1.74, 95% CI: 1.32-2.30, respectively). This was also true for concentrations in Q4 of NO_x_ (1.40, 95% CI: 1.10-1.78 and 1.38; 95% CI: 1.07- 1.77, respectively) versus Q1. When compared to Q1 of PM_2.5 absorbance_, Q4 concentrations were associated with higher unadjusted odds of ALS (1.65, 95% CI: 1.28-2.14). ^24^. The other study from the Netherlands^61^ reported higher adjusted odds of ALS per IQR increase in concentration of PM_10_ (1.10, 95% CI: 1.04-1.16), PM_coarse_ (1.06, 95% CI: 1.00-1.12), PM_2.5 absorbance_ (1.19, 95% CI: 1.10-1.28), NO_2_ (1.25, 95% CI: 1.15-1.34) and NO_x_, (1.14, 95% CI: 1.07-1.22), but not PM_2.5_ (1.05, 95% CI: 0.92-1.10). Their findings for NO_2_ and NO_x_ align with those from Seelen et al.^24^ After adjusting for age, sex, socioeconomic status and place of birth, Parks et al^60^ did not find a statistically significant association of any air pollutant examined, i.e., long-term exposure to NO_x_, elemental carbon, CO, PM_2.5_ or annual O_3_ with greater ALS risk. Similarly, Filippini et al^58^ examined exposure to PM_10_ and did not find higher odds of ALS. Among the two US studies, one^59^ did not find a significant association of any metals, pesticides, aromatic and organic/chlorinated solvents and other hazardous air pollutants. The other ^57^ reported an association of lead exposure with increased ALS risk in the states of New Hampshire/Vermont, but not in Ohio.

## Risk of Bias and Sensitivity Analyses

Using the OHAT tool, the 32 outcomes (e.g. incident PD, MS, and ALS) from the 31 papers considered for meta-analyses were assessed. None of the papers had domains with ‘Definitely high risk of bias’ and 27 had at least one domain with a ‘Probably high risk of bias’, of which 6, 1 and 1 papers had 2, 3 and 4 domains respectively (Table 6). Meta-analyses using PM as the estimator increased the magnitude of the pooled effect estimates, but statistical significance of these estimates remained unchanged from using DSL as the estimator, and therefore did not alter our conclusions. Sensitivity analysis by removing the study with the largest weight (smallest SE) was only possible for the PD outcomes (Table S8). It did not change the direction of pooled effect estimates nor statistical significance of these estimates. Meta-analyses stratified by whether studies were low risk of bias (i.e. no domains with “Probably high risk of bias”) or high risk of bias (i.e., at least one domain with “Probably high risk of bias”) was only possible for PD outcomes (Table S9). There were only a sufficient number of studies in each strata for exposures PM_10_, PM_2.5_ and NO_2_. The pooled effect estimates for PM_2.5_ in the high risk of bias category remained statistically significant.

**Table 6.** Colour-Coded list of all papers considered for meta-analyses using Risk of Bias Assessment and Level of Confidence*.

| Author: | Selection Bias | Confounding Bias | Attrition/Exclusion Bias | Detection Bias 4 | Detection Bias 5 | Selective Reporting Bias | Other Bias | Level of Confidence |
| --- | --- | --- | --- | --- | --- | --- | --- | --- |
| Cerza et al. (2018) | Probably low risk of bias | Probably low risk of bias | Probably low risk of bias | Probably low risk of bias | Probably low risk of bias | Definitely low risk of bias | Probably low risk of bias | Moderate Confidence |
| Chen et al. (2017a) | Probably low risk of bias | Probably high risk of bias | Probably low risk of bias | Probably low risk of bias | Probably low risk of bias | Definitely low risk of bias | Probably low risk of bias | Moderate Confidence |
| Kirrane et al (2015) | Probably low risk of bias | Probably low risk of bias | Probably low risk of bias | Probably low risk of bias | Probably high risk of bias | Definitely low risk of bias | Probably low risk of bias | Low Confidence |
| Lee et al. (2022) | Probably low risk of bias | Probably low risk of bias | Probably low risk of bias | Probably high risk of bias | Probably low risk of bias | Definitely low risk of bias | Probably low risk of bias | Moderate Confidence |
| Lee et al. (2016) | Probably low risk of bias | Probably high risk of bias | Probably low risk of bias | Probably high risk of bias | Definitely low risk of bias | Definitely low risk of bias | Probably low risk of bias | Moderate Confidence |
| Liu et al. (2016) | Probably low risk of bias | Probably low risk of bias | Probably low risk of bias | Probably low risk of bias | Probably low risk of bias | Definitely low risk of bias | Probably low risk of bias | Moderate Confidence |
| Palacios et al. (2014) | Probably high risk of bias | Probably low risk of bias | Probably low risk of bias | Probably low risk of bias | Probably low risk of bias | Probably low risk of bias | Probably low risk of bias | Low Confidence |
| Palacios et al. (2017) | Probably high risk of bias | Probably low risk of bias | Probably low risk of bias | Probably high risk of bias | Probably low risk of bias | Definitely low risk of bias | Probably low risk of bias | Low Confidence |
| Palacios et al. (2014) | Probably high risk of bias | Probably low risk of bias | Probably low risk of bias | Probably low risk of bias | Probably low risk of bias | Probably low risk of bias | Definitely low risk of bias | Low Confidence |
| Ritz et al. (2016) | Probably low risk of bias | Probably low risk of bias | Probably low risk of bias | Probably low risk of bias | Probably low risk of bias | Definitely low risk of bias | Probably low risk of bias | Low Confidence |
| Shin et al. (2018) | Probably low risk of bias | Probably high risk of bias | Probably low risk of bias | Probably low risk of bias | Definitely low risk of bias | Definitely low risk of bias | Probably low risk of bias | Moderate Confidence |
| Jo et al. (2021) | Probably low risk of bias | Probably high risk of bias | Probably low risk of bias | Definitely low risk of bias | Probably low risk of bias | Definitely low risk of bias | Probably low risk of bias | Moderate Confidence |
| Toro et al. (2019) | Probably low risk of bias | Probably low risk of bias | Probably low risk of bias | Probably low risk of bias | Probably low risk of bias | Definitely low risk of bias | Probably low risk of bias | Moderate Confidence |
| Willis et al. (2010) | Probably low risk of bias | Probably high risk of bias | Probably high risk of bias | Probably low risk of bias | Probably low risk of bias | Definitely low risk of bias | Probably low risk of bias | Low Confidence |
| Yu, Wei, et al. (2021) | Probably low risk of bias | Probably low risk of bias | Probably low risk of bias | Probably low risk of bias | Probably low risk of bias | Definitely low risk of bias | Probably low risk of bias | Moderate Confidence |
| Yuchi et al. (PD, 2020) | Probably low risk of bias | Probably high risk of bias | Probably low risk of bias | Probably low risk of bias | Probably low risk of bias | Probably high risk of bias | Probably low risk of bias | Low Confidence |
| Yuchi et al. (MS, 2020) | Probably low risk of bias | Probably high risk of bias | Probably low risk of bias | Probably low risk of bias | Definitely low risk of bias | Probably high risk of bias | Probably low risk of bias | Low Confidence |
| Filippini et al. (2021) | Probably high risk of bias | Probably high risk of bias | Probably low risk of bias | Definitely low risk of bias | Probably low risk of bias | Probably high risk of bias | Probably high risk of bias | Low Confidence |
| Malek et al. (2015) | Probably low risk of bias | Probably low risk of bias | Probably low risk of bias | Probably low risk of bias | Probably low risk of bias | Probably high risk of bias | Probably high risk of bias | Low Confidence |
| Parks et al. (2022) | Probably low risk of bias | Probably low risk of bias | Probably low risk of bias | Definitely low risk of bias | Definitely low risk of bias | Definitely low risk of bias | Probably low risk of bias | Moderate Confidence |
| Seelen et al. (2017) | Probably low risk of bias | Probably low risk of bias | Probably low risk of bias | Definitely low risk of bias | Probably low risk of bias | Definitely low risk of bias | Probably low risk of bias | Moderate Confidence |
| Yu, Peters, et al. (2021) | Probably low risk of bias | Probably low risk of bias | Probably low risk of bias | Probably low risk of bias | Probably low risk of bias | Probably low risk of bias | Probably low risk of bias | Moderate Confidence |
| Andrew et al. (2022) | Probably low risk of bias | Probably high risk of bias | Probably low risk of bias | Probably high risk of bias | Probably low risk of bias | Definitely low risk of bias | Probably high risk of bias | Low Confidence |
| Palacios et al. (2017) | Probably high risk of bias | Probably low risk of bias | Probably low risk of bias | Probably high risk of bias | Probably low risk of bias | Probably low risk of bias | Probably low risk of bias | Low Confidence |
| Bai et al. (2018) | Probably low risk of bias | Probably high risk of bias | Probably low risk of bias | Probably low risk of bias | Definitely low risk of bias | Definitely low risk of bias | Probably low risk of bias | Moderate Confidence |
| Chen et al. (PD, 2017b) | Definitely low risk of bias | Probably high risk of bias | Probably low risk of bias | Definitely low risk of bias | Definitely low risk of bias | Definitely low risk of bias | Probably low risk of bias | Moderate Confidence |
| Chen et al. (MS, 2017b) | Definitely low risk of bias | Probably high risk of bias | Probably low risk of bias | Definitely low risk of bias | Definitely low risk of bias | Definitely low risk of bias | Probably low risk of bias | Moderate Confidence |
| Hedstrom et al. (2023) | Probably low risk of bias | Probably low risk of bias | Probably low risk of bias | Probably low risk of bias | Probably low risk of bias | Definitely low risk of bias | Probably low risk of bias | Moderate Confidence |
| Karakis et al. (2023) | Probably high risk of bias | Probably high risk of bias | Probably low risk of bias | Probably low risk of bias | Probably low risk of bias | Definitely low risk of bias | Probably low risk of bias | Low Confidence |
| Kwon et al. (2023) | Probably low risk of bias | Probably low risk of bias | Probably low risk of bias | Probably low risk of bias | Probably low risk of bias | Definitely low risk of bias | Probably low risk of bias | Moderate Confidence |
| Rumrich et al. (2023) | Probably low risk of bias | Probably low risk of bias | Probably low risk of bias | Probably low risk of bias | Probably low risk of bias | Definitely low risk of bias | Probably low risk of bias | Moderate Confidence |
| Zhu et al. (2023) | Probably low risk of bias | Probably low risk of bias | Probably high risk of bias | Probably low risk of bias | Probably low risk of bias | Definitely low risk of bias | Probably low risk of bias | Moderate Confidence |

*\*For details about each type of bias refer to the prior published protocol*

We also performed sensitivity analyses for PD outcomes excluding studies from our meta-analysis that adjusted for comorbidities, and inferences for the meta-analyses of adjusted effect estimates was similar to the overall adjusted analysis (Table S10). We also performed sensitivity analyses excluding studies that adjusted for CVD and the adjusted effect estimates were similar as for when adjustment for comorbidities was excluded (Table S11).

The overall certainty of evidence was ‘moderate’, with the presence of unexplained inconsistency and publication bias being identified as factors that decreased confidence.

Presence of cross population and cross-study design consistency increase confidence. Therefore there is moderate confidence in the body of evidence on the association between PD risk and long-term PM2.5 exposure, but as no health effect was detected for the other exposure-outcome pairs, the level of evidence is inadequate (see Table S7).

## Discussion

In this systematic review and meta-analyses examining the association of long-term outdoor air pollution exposure with three neurodegenerative diseases PD, MS and MND, most studies were from North America and Europe and assessed PD incidence. The meta-analysis of incident PD studies provides evidence for an association of long-term exposure to ambient PM_2.5_. By contrast, there were no associations of PM_10_, PM_2.5-10_, NO_2_, NO_x_ or annual O_3_ with PD incidence. Among the exposure outcome pairs with sufficient number of studies, there was evidence of small study effect (i.e., an indicator of publication bias) for the association of PM_2.5_ and PM_10_, but not NO_2_ with PD incidence. To mitigate risk of small study effects in future research, focus can be on larger sample sizes to strengthen evidence through biobank or registry-based studies.

There have been several systematic reviews and meta-analyses on the association of long-term outdoor air pollution exposure with PD incidence, but they vary in selection of pollutants, number of databases searched and analytical approaches (e.g., subgroup analyses and sensitivity analyses, risk of bias assessment and certainty of evidence assessment).^25–30^ Two only included PM and of the ones that included gaseous pollutants, none differentiated between annual and seasonal O_3_. The latter is important because O_3_ levels fluctuate throughout the year. The maximum number of databases searched was six (four academic and two World Health Organisation grey literature databases)^25^, which is fewer than the eight databases searched in this systematic review. Only two carried out subgroup analyses, using different study and individual characteristics to stratify groups, and none carried out certainty of evidence assessment. Of note, two meta-analyses identified greater PD incidence with exposure to PM_2.5_ similar to our study, but the effect estimates in those studies were higher (1.34, 95% CI: 1.04, 1.73) and lower (1.01, 95% CI: 1.01-1.02) than reported here.^26,29^ Two identified a statistically significant association of CO with incident PD and four^25,27–29^ of O_3_ exposure, but meta-analyses for each pollutant were heavily weighted by one study published by Lee et al^62^ (40-55% and 88-92%, respectively). Dhiman et al^25^ noted that once they removed Lee et al^62^ from their meta-analysis, the association between PD risk and NO_2_ diminished. Additionally, Hu et al^28^ and Fu et al^26^ used overlapping data from two articles based on the same population and therefore pooled effect estimates may artificially exaggerate the weight of specific studies. Only Han et al^27^ performed a risk of bias assessment, and none included a certainty of evidence assessment.

For MS, our meta-analysis did not identify an association of long-term exposure to PM_2.5_ or NO_2_ with risk of disease, but two prior meta-analyses did. Lofti et al^31^ found in their meta-analysis of three studies an increased risk of MS relapse and incidence from PM_2.5_ and PM_10_ exposure. Tang et al^32^ identified an association of PM_10_ exposure, but not with other particulate or gaseous pollutants. These meta-analyses had different inclusion criteria, defining outcome as incidence and relapse, and included exposure windows of less than one year.

Finally, this is the first systematic review on the association of air pollution exposure to MND incidence, identifying six primary research studies that provided effect estimates on pesticides, organic/chloride solvents, particulate, gaseous and hazardous air pollutant exposure.

### Biological plausibility

Evidence from animal and human controlled studies demonstrate that oxidative stress and neuroinflammation play an important role in the abnormal aggregation of α-synuclein and degeneration of the dopaminergic neurons, characteristics of PD.^63,64^ Another study suggested that air pollution exposure and polymorphisms in the proinflammatory cytokine gene interleukin-1β facilitate the risk of developing PD. ^48^ Some have also hypothesized that PM_2.5_ produces neuroinflammation leading to altered innate immune responses in anatomical areas of the brain and enhancement of α-synuclein aggregation. ^17^ Variability in effect estimates in prior meta-analyses may be related to different composition of PM_2.5_, as Finkelstein and Jerrett^65^ observed an association between ambient levels of airborne manganese (which make up PM_2.5_) and PD risk. Proposed pathological mechanisms also implicate CO as it bonds with haemoglobin to form carbon-oxyhaemoglobin, reducing transport of oxygen that is required to form levodopa, a precursor of dopamine. ^66^ The multifactorial aetiology of PD makes understanding the association between pollutant and PD risk difficult to unpick.

The pro-inflammatory role of air pollution is proposed to play a key role in the risk of MS too, through upregulating expression of CCR6 on circulating CD4+ T cells and inducing innate immune cells to produce Th17 polarising cytokines.^67^ Relapses have been reported to be more frequent following exposure to high levels of PM_10_ in MS patients not using immunomodulatory drugs (e.g., β-interferon), but no effect on those who were.^68^ Proposed indirect effects of air pollution on MS risk include infection risk and Vitamin D production. Different infections (e.g., Epstein-Barr virus) have been associated with MS risk, and air pollution may enhance susceptibility to infections.^68–70^ Low levels of Vitamin D, an immunomodulatory agent whose production is dependent on ultraviolet B (UVB) exposure, is associated with greater risk of MS relapse.^71^ Ambient air pollution can absorb UVB in the atmosphere, reducing ground level exposure leading to a reduction in Vitamin D production and consequently greater risk of disease relapse.^70,72,73^

There was a positive association between ALS and several traffic-related air pollutants (PM_2.5 absorbance_, NO_2_ and NO_x_), which aligns with findings on ALS risk in occupational studies for truck and bus drivers.^14,74,75^ Traffic-related pollution may contain elements characterised by neurotoxic effects, e.g., lead, cadmium, and mercury, but their association with ALS is inconclusive.^76,77^ Toxicological and animal studies provide evidence of biological pathways leading to neurodegenerative diseases, through neuroinflammation and oxidative stress, but these pathways are not specific to MND.^24^ Although large-scale epidemiologic studies can assess the association more robustly, this approach is limited by low incidence rates of disease groups with distinct clinical and pathological phenotypes.

### Strengths

There are several strengths in our systematic review and meta-analysis. To date, the search strategy in this study included the largest number of databases for all three disease outcomes and is the first systematic review for MND. The methods follow the gold-standard practice of using two independent reviewers for each stage of screening and assessment (e.g., screening, data extraction, risk of bias assessments). ^78^ Furthermore, outcome misclassification was reduced through our inclusion criteria requiring confirmation by a medical professional and that exposure preceded the outcome. Our exposure-outcome pairs were analysed separately by adjusted and unadjusted effect estimates and we differentiated between warm season and annual O3. Effect estimates from the most restrictive analysis models (e.g., adjustment for lifestyle factors such as smoking, alcohol consumption, diet, and physical activity, indoor environmental factors, versus not) were selected. Air pollution studies have substantial heterogeneity, due to differences in study designs, population size and composition and exposure assessment methods, and those assessing rare outcomes are likely to have small sample sizes. By using PM estimator, in addition to DSL, our sensitivity analyses provide more reliable estimates of between study variance and heterogeneity.

## Limitations

Despite unique strengths of this study, there are limitations in this systematic review and meta-analysis. Firstly, more than half of the included studies were based on a case-control design, which limits ability to infer causal relationships. Secondly, statistical adjustment was required for case control and cohort studies and adjustments differed between primary studies which may be a source of high between study heterogeneity (i.e., heterogeneity was low for unadjusted and high for adjusted in meta-analyses for PM_2.5_ and PD). Thirdly, most primary studies were carried out in high-income countries, which tend to have lower levels of population exposure to air pollution than low- and middle-income countries (LMIC) and this greatly limits the generalisability of our results.^79^ This is a difficult balance however as incidence/prevalence of neurodegenerative diseases such as PD, MS and MND are likely to be underreported in LMIC due to geographical and financial barriers to healthcare services.^80,81^ Fourth, methodological limitations in primary studies, such as exposure misclassification due to scope of exposure assessments could have affected our results. Fifth, we were not able to precisely assess the duration of exposure to examine whether our definition of long-term exposure (1 or more years) is enough time to develop neurodegenerative movement diseases in association with air pollution exposure.

## Conclusion

This systematic review identifies growing evidence on the association of outdoor air pollution with incidence of neurodegenerative diseases. There is evidence on the association of PD risk with long-term exposure to PM_2.5_, but no association identified with other pollutants. No association was identified, or assessed, between pollutant exposure and MS and MND incidence due to a paucity of studies with clear epidemiological and clinical definitions of outcomes. Although large-scale epidemiologic studies can assess the association more robustly, this approach is limited by low incidence rates of disease groups with distinct clinical and pathological phenotypes.

## Appendix

Supplementary Tables and Figures

Submitted PROSPERO Protocol: Registration number is CRD42023417961

## SUPPLEMENTARY MATERIALS

S1: List of excluded studies and reason for exclusion at full text screening stage

https://docs.google.com/spreadsheets/d/1X06zVqINJ7QZLZ7JMm7AnNCZDqeNh2KgSyWi yuqx4fE/edit?gid=1052957813#gid=105295781

S2: Data Extraction Form https://docs.google.com/spreadsheets/d/1kmv5NwF8vATMpBir_5ZVVkCr7BWZq60N/edit? gid=1016733053#gid=1016733053

S3: Complete table of extracted data points

https://docs.google.com/spreadsheets/d/1E98kaFxheJ1K5WpCAcYwjgMuVUB7-HrmZdfvXc8WJtg/edit?usp=sharing

## Data Availability

All data produced in the present study are available upon reasonable request to the authors

