## Supplemental Tables and Figures for "Association of Long-Term Outdoor Air Pollution Exposure with Incidence of Parkinson’s Disease, Multiple Sclerosis and Motor Neuron Diseases: A Systematic Review and Meta-Analysis"

**Supplementary Tables:**

**Table S1**. Deviations from Original Protocol

| Aspect of Original Protocol | Deviation |
|---|---|
| 4.6.2 Minimum number of studies required for meta-analysis | The published protocol stated, "when four or more independent studies reporting on an exposure-outcome pair are available, we will meta-analyze the associations." This was reduced to three or more studies in the final paper. |
| 4.6.3 Use of the REML estimator | The published protocol stated that "in sensitivity analysis, we will also use the Restricted Maximum Likelihood estimator (REML)." In light of more recent publications on estimator performance, we chose to use the Paule-Mandel in sensitivity analysis instead of the REML. |
| 4.6.4 Sensitivity Analysis | The published protocol stated that a sensitivity analysis would be run excluding studies with a high risk of bias, defined as studies with at least one domain with a "definitely high risk of bias" rating. There were no studies that fulfilled this criterion and therefore this sensitivity analysis was not carried out. We were only able to carry out sensitivity analysis comparing combined "Definitely low risk of bias" and "Probably low risk of bias", versus "Probably high risk of bias". For long-term exposure from $PM_{2.5}$, $PM_{10}$ and $NO_2$ with Parkinson's Disease as the outcome and effect estimates were adjusted. |
| 4.6.5 Subgroup Analysis | Due to a lack of studies, we were unable to conduct subgroup analysis by sex, age, and ethnicity. |
| 4.7 ERF | Due to a limited number of studies which recorded categorical results for each pollutant, we were only able to include ERFs for PD outcomes and $PM_{2.5}$, $PM_{10}$ and $NO_2$ pollutants. |

**Table S2**. Database Search Terms

| Database | Search Terms |
|---|---|
| Medline via Ovid | (dementia* or alzheimer* or "anterior horn disease" or "amyotrophic lateral sclerosis" or demyelinati* or "disseminated sclerosis" or encephalomyelitis or parkinson* or "lewy bod*" or "motor neuron disease" or "multiple sclerosis" or neurodegenerat* or "neuro-degenerat*" or parkinson* or "primary lateral sclerosis" or "progressive bulbar palsy" or "progressive muscular atrophy" or "progressive spinal muscl*" or "pseudobulbar palsy" or "spinal muscular atrophy").ti,ab,kw. or exp Neurodegenerative Diseases/ or exp dementia/<br><br>AND |

| | |
|---|---|
| | ("Air pollut*" or "air quality" or "ambient particulate matter" or "black carbon" or "black smoke" or "carbon dioxide" or "carbon monoxide" or "carbonic oxide" or "Coarse partic*" or diesel or "elemental carbon" or "environmental exposure" or "environmental pollution" or "exhaust gas" or "exhaust fume" or "exhaust emission*" or "fine partic*" or hydrocarbons or (Indust* adj2 pollut*) or "inhalable particle" or nitrates or "nitric oxide" or "nitrogen dioxide" or "nitrogen oxide*" or "nitrous oxide" or ozone or Particle* or particulate* or "Peroxyacetyl Nitrate" or pollut* or "Polycyclic Aromatic Hydrocarbons" or ((proximity or distance) adj3 (road* or highway* or street* or motorway* or "point source*")) or smog or Soot or "Sulfur dioxide" or "Sulfur Monoxide" or "Sulfur Oxides" or "sulphur dioxide" or "sulphur monoxide" or "sulphur oxides" or traffic or Ultrafine or "Ultra-fine" or "vehicle emissions" or "volatile Organic Compound*").ti,ab,kw. or air pollution/ or Air Pollutants/ or Vehicle Emissions/ or Particulate Matter/ or smog/ or soot/ or carbon dioxide/ or carbon monoxide/ or nitric oxide/ or environmental exposure/ or hydrocarbons/ or nitrates/ or nitrogen dioxide/ or nitrogen oxides/ or ozone/ or polycyclic Aromatic Hydrocarbons/ or sulfur dioxide/ or sulfur oxides/ or vehicle emissions/ or volatile organic compounds/<br><br>AND<br><br>(cohort or "ecological stud*" or "cross sectional" or longitudinal* or observational or association or prospective or retrospective or "case control").ti,ab,kw. or exp cohort studies/ or exp case-control studies/ or cross-sectional studies/ or follow-up studies/ or longitudinal studies/ or observational study/ or cluster analysis/ or geographical mapping/ |
| Embase (via Ovid) | (dementia* or alzheimer* or "anterior horn disease" or "amyotrophic lateral sclerosis" or demyelinati* or "disseminated sclerosis" or encephalomyelitis or parkinson* or "lewy bod*" or "motor neuron disease" or "multiple sclerosis" or neurodegenerat* or "neuro-degenerat*" or parkinson* or "primary lateral sclerosis" or "progressive bulbar palsy" or "progressive muscular atrophy" or "progressive spinal muscl*" or "pseudobulbar palsy" or "spinal muscular atrophy").ti,ab,kw. or exp Neurodegenerative Diseases/ or exp dementia/<br><br>AND<br><br>("Air pollut*" or "air quality" or "ambient particulate matter" or "black carbon" or "black smoke" or "carbon dioxide" or "carbon monoxide" or "carbonic oxide" or "Coarse partic*" or diesel or "elemental carbon" or "environmental exposure" or "environmental pollution" or "exhaust gas" or "exhaust fume" or "exhaust emission*" or "fine partic*" or hydrocarbons or (Indust* adj2 pollut*) or "inhalable particle" or nitrates or "nitric oxide" or "nitrogen dioxide" or "nitrogen oxide*" or "nitrous oxide" or ozone or Particle* or particulate* or "Peroxyacetyl Nitrate" or pollut* or "Polycyclic Aromatic Hydrocarbons" or ((proximity or distance) adj3 (road* or highway* or street* or motorway* or "point source*")) or smog or Soot or "Sulfur dioxide" or "Sulfur Monoxide" or "Sulfur Oxides" or "sulphur dioxide" or "sulphur monoxide" or "sulphur |

| | |
|---|---|
| | oxides" or traffic or Ultrafine or "Ultra-fine" or "vehicle emissions" or "volatile Organic Compound*").ti,ab,kw. Or air pollution/ OR Air Pollutant/ or air quality/ or black carbon/ or diesel particulate matter/ or exhaust gas/ or hydrocarbon/ or Particulate Matter/ OR smog/ OR soot/ OR carbon dioxide/ or carbon monoxide/ OR nitric oxide/ OR environmental exposure/ OR nitrates/ OR nitrogen dioxide/ OR nitrogen oxide/ OR nitrous oxide/ OR ozone/ OR polycyclic Aromatic Hydrocarbons/ OR sulfur dioxide/ OR sulfur oxides/ OR volatile organic compounds/ OR smog/ OR soot/ OR traffic/ OR environmental surveillance/<br><br>AND<br><br>(cohort or "ecological stud*" or "cross sectional" or longitudinal* or observational or association or prospective or retrospective or "case control").ti,ab,kw. OR exp cohort studies/ OR exp case-control studies/ OR cross-sectional studies/ OR follow-up studies/ OR longitudinal studies/ OR observational study/ OR evaluation study/ OR program evaluation/ OR regression analysis/ or or cluster analysis/ or geographical mapping/) |
| Cochrane Library | (dementia* or alzheimer* or "anterior horn disease" or "amyotrophic lateral sclerosis" or demyelinati* or "disseminated sclerosis" or encephalomyelitis or parkinson* or "lewy bod*" or "motor neuron disease" or "multiple sclerosis" or neurodegenerat* or "neuro-degenerat*" or parkinson* or "primary lateral sclerosis" or "progressive bulbar palsy" or "progressive muscular atrophy" or "progressive spinal muscl*" or "pseudobulbar palsy" or "spinal muscular atrophy"):ti,ab,kw or MeSH descriptor: [Neurodegenerative Diseases] explode all trees or MeSH descriptor: [dementia] explode all trees<br><br>AND<br><br>("Air pollut*" or "air quality" or "ambient particulate matter" or "black carbon" or "black smoke" or "carbon dioxide" or "carbon monoxide" or "carbonic oxide" or "Coarse partic*" or diesel or "elemental carbon" or "environmental exposure" or "environmental pollution" or "exhaust gas" or "exhaust fume" or "exhaust emission*" or "fine partic*" or hydrocarbons or (Indust* adj2 pollut*) or "inhalable particle" or nitrates or "nitric oxide" or "nitrogen dioxide" or "nitrogen oxide*" or "nitrous oxide" or ozone or Particle* or particulate* or "Peroxyacetyl Nitrate" or pollut* or "Polycyclic Aromatic Hydrocarbons" or ((proximity or distance) adj3 (road* or highway* or street* or motorway* or "point source*")) or smog or Soot or "Sulfur dioxide" or "Sulfur Monoxide" or "Sulfur Oxides" or "sulphur dioxide" or "sulphur monoxide" or "sulphur oxides" or traffic or Ultrafine or "Ultra-fine" or "vehicle emissions" or "volatile Organic Compound*");ti,ab,kw or MeSH descriptor: [air pollution] this term only or MeSH descriptor: [Air Pollutants] this term only or MeSH descriptor: [Vehicle Emissions] this term only or MeSH descriptor: [Particulate Matter] this term only or MeSH descriptor: [smog] this term only or MeSH descriptor: [soot] this term only or MeSH descriptor: [carbon dioxide] this term only or MeSH descriptor: [carbon monoxide] this term only or MeSH |

| | |
|---|---|
| | descriptor: [nitric oxide] this term only or MeSH descriptor: [environmental exposure] this term only or MeSH descriptor: [hydrocarbons] this term only or MeSH descriptor: [nitrates] this term only or MeSH descriptor: [nitrogen dioxide] this term only or MeSH descriptor: [nitrogen oxides] this term only or MeSH descriptor: [ozone] this term only or MeSH descriptor: [polycyclic Aromatic Hydrocarbons] this term only or MeSH descriptor: [sulfur dioxide] this term only or MeSH descriptor: [sulfur oxides] this term only or MeSH descriptor: [vehicle emissions] this term only or MeSH descriptor: [volatile organic compounds] this term only<br><br>AND<br><br>(cohort or "ecological stud*" or "cross sectional" or longitudinal* or observational or association or prospective or retrospective or "case control").ti,ab,kw. or MeSH descriptor: [cohort studies] explode all trees or MeSH descriptor: [case-control studies] explode all trees or MeSH descriptor: [cross-sectional studies] this term only or MeSH descriptor: [follow-up studies] this term only or MeSH descriptor: [longitudinal studies/ or MeSH descriptor: [observational study] this term only or MeSH descriptor: [cluster analysis] this term only or MeSH descriptor: [geographical mapping] this term only |
| PsycINFO (via Ebscohost) | TI (dementia* or alzheimer* or "anterior horn disease" or "amyotrophic lateral sclerosis" or demyelinati* or "disseminated sclerosis" or encephalomyelitis or parkinson* or "lewy bod*" or "motor neuron disease" or "multiple sclerosis" or neurodegenerat* or "neuro-degenerat*" or parkinson* or "primary lateral sclerosis" or "progressive bulbar palsy" or "progressive muscular atrophy" or "progressive spinal muscl*" or "pseudobulbar palsy" or "spinal muscular atrophy") OR AB (dementia* or alzheimer* or "anterior horn disease" or "amyotrophic lateral sclerosis" or demyelinati* or "disseminated sclerosis" or encephalomyelitis or parkinson* or "lewy bod*" or "motor neuron disease" or "multiple sclerosis" or neurodegenerat* or "neuro-degenerat*" or parkinson* or "primary lateral sclerosis" or "progressive bulbar palsy" or "progressive muscular atrophy" or "progressive spinal muscl*" or "pseudobulbar palsy" or "spinal muscular atrophy") OR KW (dementia* or alzheimer* or "anterior horn disease" or "amyotrophic lateral sclerosis" or demyelinati* or "disseminated sclerosis" or encephalomyelitis or parkinson* or "lewy bod*" or "motor neuron disease" or "multiple sclerosis" or neurodegenerat* or "neuro-degenerat*" or parkinson* or "primary lateral sclerosis" or "progressive bulbar palsy" or "progressive muscular atrophy" or "progressive spinal muscl*" or "pseudobulbar palsy" or "spinal muscular atrophy") OR DE "Neurodegenerative Diseases" OR DE "Alzheimer's Disease" OR DE "Amyotrophic Lateral Sclerosis" OR DE "Dementia with Lewy Bodies" OR DE "Parkinson's Disease" " OR DE “Dementia”<br><br>AND |

| | |
|---|---|
| | TI ("Air pollut*" or "air quality" or "ambient particulate matter" or "black carbon" or "black smoke" or "carbon dioxide" or "carbon monoxide" or "carbonic oxide" or "Coarse partic*" or diesel or "elemental carbon" or "environmental exposure" or "environmental pollution" or "exhaust gas" or "exhaust fume" or "exhaust emission*" or "fine partic*" or hydrocarbons or (Indust* N2 pollut*) or "inhalable particle" or nitrates or "nitric oxide" or "nitrogen dioxide" or "nitrogen oxide*" or "nitrous oxide" or ozone or Particle* or particulate* or "Peroxyacetyl Nitrate" or pollut* or "Polycyclic Aromatic Hydrocarbons" or ((proximity or distance) N3 (road* or highway* or street* or motorway* or "point source*")) or smog or Soot or "Sulfur dioxide" or "Sulfur Monoxide" or "Sulfur Oxides" or "sulphur dioxide" or "sulphur monoxide" or "sulphur oxides" or traffic or Ultrafine or "Ultra-fine" or "vehicle emissions" or "volatile Organic Compound*") OR AB ("Air pollut*" or "air quality" or "ambient particulate matter" or "black carbon" or "black smoke" or "carbon dioxide" or "carbon monoxide" or "carbonic oxide" or "Coarse partic*" or diesel or "elemental carbon" or "environmental exposure" or "environmental pollution" or "exhaust gas" or "exhaust fume" or "exhaust emission*" or "fine partic*" or hydrocarbons or (Indust* N2 pollut*) or "inhalable particle" or nitrates or "nitric oxide" or "nitrogen dioxide" or "nitrogen oxide*" or "nitrous oxide" or ozone or Particle* or particulate* or "Peroxyacetyl Nitrate" or pollut* or "Polycyclic Aromatic Hydrocarbons" or ((proximity or distance) N3 (road* or highway* or street* or motorway* or "point source*")) or smog or Soot or "Sulfur dioxide" or "Sulfur Monoxide" or "Sulfur Oxides" or "sulphur dioxide" or "sulphur monoxide" or "sulphur oxides" or traffic or Ultrafine or "Ultra-fine" or "vehicle emissions" or "volatile Organic Compound*") OR KW ("Air pollut*" or "air quality" or "ambient particulate matter" or "black carbon" or "black smoke" or "carbon dioxide" or "carbon monoxide" or "carbonic oxide" or "Coarse partic*" or diesel or "elemental carbon" or "environmental exposure" or "environmental pollution" or "exhaust gas" or "exhaust fume" or "exhaust emission*" or "fine partic*" or hydrocarbons or (Indust* N2 pollut*) or "inhalable particle" or nitrates or "nitric oxide" or "nitrogen dioxide" or "nitrogen oxide*" or "nitrous oxide" or ozone or Particle* or particulate* or "Peroxyacetyl Nitrate" or pollut* or "Polycyclic Aromatic Hydrocarbons" or ((proximity or distance) N3 (road* or highway* or street* or motorway* or "point source*")) or smog or Soot or "Sulfur dioxide" or "Sulfur Monoxide" or "Sulfur Oxides" or "sulphur dioxide" or "sulphur monoxide" or "sulphur oxides" or traffic or Ultrafine or "Ultra-fine" or "vehicle emissions" or "volatile Organic Compound*") OR DE "Carbon Dioxide" OR DE "Carbon Monoxide" OR DE "Nitric Oxide" OR DE "Pollution"<br><br>AND<br><br>TI (cohort or "ecological stud*" or "cross sectional" or longitudinal* or observational or association or prospective or retrospective or "case control") OR AB (cohort or "ecological stud*" or "cross sectional" or longitudinal* or observational or association or prospective or retrospective or "case control") OR KW (cohort or "ecological stud*" or "cross sectional" or longitudinal* or observational or association or prospective or retrospective |

| | |
|---|---|
| | or "case control") OR DE "Longitudinal Studies" OR DE "Prospective Studies" OR DE "Retrospective Studies" |
| Cinahl (via Ebscohost) | TI (dementia* or alzheimer* or "anterior horn disease" or "amyotrophic lateral sclerosis" or demyelinati* or "disseminated sclerosis" or encephalomyelitis or parkinson* or "lewy bod*" or "motor neuron disease" or "multiple sclerosis" or neurodegenerat* or "neuro-degenerat*" or parkinson* or "primary lateral sclerosis" or "progressive bulbar palsy" or "progressive muscular atrophy" or "progressive spinal muscl*" or "pseudobulbar palsy" or "spinal muscular atrophy") OR AB (dementia* or alzheimer* or "anterior horn disease" or "amyotrophic lateral sclerosis" or demyelinati* or "disseminated sclerosis" or encephalomyelitis or parkinson* or "lewy bod*" or "motor neuron disease" or "multiple sclerosis" or neurodegenerat* or "neuro-degenerat*" or parkinson* or "primary lateral sclerosis" or "progressive bulbar palsy" or "progressive muscular atrophy" or "progressive spinal muscl*" or "pseudobulbar palsy" or "spinal muscular atrophy") OR (MH "Neurodegenerative Diseases+") OR (MH "Dementia")<br><br>AND<br><br>TI ("Air pollut*" or "air quality" or "ambient particulate matter" or "black carbon" or "black smoke" or "carbon dioxide" or "carbon monoxide" or "carbonic oxide" or "Coarse partic*" or diesel or "elemental carbon" or "environmental exposure" or "environmental pollution" or "exhaust gas" or "exhaust fume" or "exhaust emission*" or "fine partic*" or hydrocarbons or (Indust* N2 pollut*) or "inhalable particle" or nitrates or "nitric oxide" or "nitrogen dioxide" or "nitrogen oxide*" or "nitrous oxide" or ozone or Particle* or particulate* or "Peroxyacetyl Nitrate" or pollut* or "Polycyclic Aromatic Hydrocarbons" or ((proximity or distance) N3 (road* or highway* or street* or motorway* or "point source*")) or smog or Soot or "Sulfur dioxide" or "Sulfur Monoxide" or "Sulfur Oxides" or "sulphur dioxide" or "sulphur monoxide" or "sulphur oxides" or traffic or Ultrafine or "Ultra-fine" or "vehicle emissions" or "volatile Organic Compound*") OR AB ("Air pollut*" or "air quality" or "ambient particulate matter" or "black carbon" or "black smoke" or "carbon dioxide" or "carbon monoxide" or "carbonic oxide" or "Coarse partic*" or diesel or "elemental carbon" or "environmental exposure" or "environmental pollution" or "exhaust gas" or "exhaust fume" or "exhaust emission*" or "fine partic*" or hydrocarbons or (Indust* N2 pollut*) or "inhalable particle" or nitrates or "nitric oxide" or "nitrogen dioxide" or "nitrogen oxide*" or "nitrous oxide" or ozone or Particle* or particulate* or "Peroxyacetyl Nitrate" or pollut* or "Polycyclic Aromatic Hydrocarbons" or ((proximity or distance) N3 (road* or highway* or street* or motorway* or "point source*")) or smog or Soot or "Sulfur dioxide" or "Sulfur Monoxide" or "Sulfur Oxides" or "sulphur dioxide" or "sulphur monoxide" or "sulphur oxides" or traffic or Ultrafine or "Ultra-fine" or "vehicle emissions" or "volatile Organic Compound*") OR (MH "Air Pollution") OR (MH "Air Pollutants") OR (MH "Carbon Dioxide") OR (MH "Carbon Monoxide") OR (MH "Environmental Exposure") OR (MH "Environmental Pollution") OR (MH "Hydrocarbons") OR (MH "Nitrates") OR |

| | |
|---|---|
| | (MH "Nitric Oxide") OR (MH "Nitrogen Oxides") OR (MH "Nitrous Oxide") OR (MH "Ozone") OR (MH "Smog") OR (MH "Traffic Pollution") OR (MH "Particulate Matter") OR (MH "Motor Vehicle Emissions")<br><br>AND<br><br>TI (cohort or "ecological stud*" or "cross sectional" or longitudinal* or observational or association or prospective or retrospective or "case control") OR AB (cohort or "ecological stud*" or "cross sectional" or longitudinal* or observational or association or prospective or retrospective or "case control") OR (MH "Cross Sectional Studies") OR (MH "Prospective Studies") |
| Global Health (via Ebscohost) | TI (dementia* or alzheimer* or "anterior horn disease" or "amyotrophic lateral sclerosis" or demyelinati* or "disseminated sclerosis" or encephalomyelitis or parkinson* or "lewy bod*" or "motor neuron disease" or "multiple sclerosis" or neurodegenerat* or "neuro-degenerat*" or parkinson* or "primary lateral sclerosis" or "progressive bulbar palsy" or "progressive muscular atrophy" or "progressive spinal muscl*" or "pseudobulbar palsy" or "spinal muscular atrophy") OR AB (dementia* or alzheimer* or "anterior horn disease" or "amyotrophic lateral sclerosis" or demyelinati* or "disseminated sclerosis" or encephalomyelitis or parkinson* or "lewy bod*" or "motor neuron disease" or "multiple sclerosis" or neurodegenerat* or "neuro-degenerat*" or parkinson* or "primary lateral sclerosis" or "progressive bulbar palsy" or "progressive muscular atrophy" or "progressive spinal muscl*" or "pseudobulbar palsy" or "spinal muscular atrophy") OR (DE "dementia") OR (DE "Alzheimer's disease") OR (DE "Parkinson's disease") OR (DE "multiple sclerosis")<br><br>AND<br><br>TI ("Air pollut*" or "air quality" or "ambient particulate matter" or "black carbon" or "black smoke" or "carbon dioxide" or "carbon monoxide" or "carbonic oxide" or "Coarse partic*" or diesel or "elemental carbon" or "environmental exposure" or "environmental pollution" or "exhaust gas" or "exhaust fume" or "exhaust emission*" or "fine partic*" or hydrocarbons or (Indust* N2 pollut*) or "inhalable particle" or nitrates or "nitric oxide" or "nitrogen dioxide" or "nitrogen oxide*" or "nitrous oxide" or ozone or Particle* or particulate* or "Peroxyacetyl Nitrate" or pollut* or "Polycyclic Aromatic Hydrocarbons" or ((proximity or distance) N3 (road* or highway* or street* or motorway* or "point source*")) or smog or Soot or "Sulfur dioxide" or "Sulfur Monoxide" or "Sulfur Oxides" or "sulphur dioxide" or "sulphur monoxide" or "sulphur oxides" or traffic or Ultrafine or "Ultra-fine" or "vehicle emissions" or "volatile Organic Compound*") OR AB ("Air pollut*" or "air quality" or "ambient particulate matter" or "black carbon" or "black smoke" or "carbon dioxide" or "carbon monoxide" or "carbonic oxide" or "Coarse partic*" or diesel or "elemental carbon" or "environmental exposure" or "environmental pollution" or "exhaust gas" or "exhaust fume" or "exhaust emission*" or "fine partic*" or hydrocarbons or (Indust* N2 pollut*) or "inhalable particle" or |

| | |
|---|---|
| | nitrates or "nitric oxide" or "nitrogen dioxide" or "nitrogen oxide*" or "nitrous oxide" or ozone or Particle* or particulate* or "Peroxyacetyl Nitrate" or pollut* or "Polycyclic Aromatic Hydrocarbons" or ((proximity or distance) N3 (road* or highway* or street* or motorway* or "point source*")) or smog or Soot or "Sulfur dioxide" or "Sulfur Monoxide" or "Sulfur Oxides" or "sulphur dioxide" or "sulphur monoxide" or "sulphur oxides" or traffic or Ultrafine or "Ultra-fine" or "vehicle emissions" or "volatile Organic Compound*") OR (DE "air pollution") OR DE ("air quality") OR (DE "carbon dioxide")) OR (DE "carbon monoxide") OR DE ("exhaust gases") OR (DE "pollution") OR (DE "hydrocarbons") OR (DE "nitrates") OR (DE "nitrous oxide") OR (DE "nitrogen dioxide") OR DE ("nitrogen oxides") OR (DE "ozone") OR (DE "sulfur dioxide")<br><br>AND<br><br>TI (cohort or "ecological stud*" or "cross sectional" or longitudinal* or observational or association or prospective or retrospective or "case control") OR AB (cohort or "ecological stud*" or "cross sectional" or longitudinal* or observational or association or prospective or retrospective or "case control") OR DE "longitudinal studies" |
| Web of Science (Core Collection) | TS (dementia* or alzheimer* or "anterior horn disease" or "amyotrophic lateral sclerosis" or demyelinati* or "disseminated sclerosis" or encephalomyelitis or parkinson* or "lewy bod*" or "motor neuron disease" or "multiple sclerosis" or neurodegenerat* or "neuro-degenerat*" or parkinson* or "primary lateral sclerosis" or "progressive bulbar palsy" or "progressive muscular atrophy" or "progressive spinal muscl*" or "pseudobulbar palsy" or "spinal muscular atrophy")<br><br>AND<br><br>TS ("Air pollut*" or "air quality" or "ambient particulate matter" or "black carbon" or "black smoke" or "carbon dioxide" or "carbon monoxide" or "carbonic oxide" or "Coarse partic*" or diesel or "elemental carbon" or "environmental exposure" or "environmental pollution" or "exhaust gas" or "exhaust fume" or "exhaust emission*" or "fine partic*" or hydrocarbons or (Indust* Near/2 pollut*) or "inhalable particle" or nitrates or "nitric oxide" or "nitrogen dioxide" or "nitrogen oxide*" or "nitrous oxide" or ozone or Particle* or particulate* or "Peroxyacetyl Nitrate" or pollut* or "Polycyclic Aromatic Hydrocarbons" or ((proximity or distance) Near/3 (road* or highway* or street* or motorway* or "point source*")) or smog or Soot or "Sulfur dioxide" or "Sulfur Monoxide" or "Sulfur Oxides" or "sulphur dioxide" or "sulphur monoxide" or "sulphur oxides" or traffic or Ultrafine or "Ultra-fine" or "vehicle emissions" or "volatile Organic Compound*")<br><br>AND |

| | |
|---|---|
| | TS (cohort or "ecological stud*" or "cross sectional" or longitudinal* or observational or association or prospective or retrospective or "case control") |
| Scopus | Title-Abs-Key (dementia* or alzheimer* or "anterior horn disease" or "amyotrophic lateral sclerosis" or demyelinati* or "disseminated sclerosis" or encephalomyelitis or parkinson* or "lewy bod*" or "motor neuron disease" or "multiple sclerosis" or neurodegenerat* or "neuro-degenerat*" or parkinson* or "primary lateral sclerosis" or "progressive bulbar palsy" or "progressive muscular atrophy" or "progressive spinal muscl*" or "pseudobulbar palsy" or "spinal muscular atrophy")<br><br>AND<br><br>Title-Abs-Key ("Air pollut*" or "air quality" or "ambient particulate matter" or "black carbon" or "black smoke" or "carbon dioxide" or "carbon monoxide" or "carbonic oxide" or "Coarse partic*" or diesel or "elemental carbon" or "environmental exposure" or "environmental pollution" or "exhaust gas" or "exhaust fume" or "exhaust emission*" or "fine partic*" or hydrocarbons or (Indust* W/2 pollut*) or "inhalable particle" or nitrates or "nitric oxide" or "nitrogen dioxide" or "nitrogen oxide*" or "nitrous oxide" or ozone or Particle* or particulate* or "Peroxyacetyl Nitrate" or pollut* or "Polycyclic Aromatic Hydrocarbons" or ((proximity or distance) W/3 (road* or highway* or street* or motorway* or "point source*")) or smog or Soot or "Sulfur dioxide" or "Sulfur Monoxide" or "Sulfur Oxides" or "sulphur dioxide" or "sulphur monoxide" or "sulphur oxides" or traffic or Ultrafine or "Ultra-fine" or "vehicle emissions" or "volatile Organic Compound*")<br><br>AND<br><br>Title-Abs-Key (cohort or "ecological stud*" or "cross sectional" or longitudinal* or observational or association or prospective or retrospective or "case control") |

**Table S3**. Variables extracted in Data Extraction.

- Publication year
- authors
- funding source
- Study country and name
- outcome
- air pollutant, mean, median, min, max, IQR concentrations.
- study design
- study population
- population size
- incident cases
- ethnicity
- proportion female
- mean, min, max age
- age onset
- study start and end, and follow up value, and unit,
- outcome diagnosis, ascertainment, and definition
- exposure assessment method, its spatial resolution and spatial scale
- exposure duration
- statistical model
- unadjusted hr, or, or hr
- adjusted RR, HR, or OR
    - confounders adjusted for: age, sex, family history, bmi, smoking, caffeine, diet, alcohol, activity, apoe4, education, socioeconomic indicators, urban vs, rural, comorbidities
- p values
- Subgroup and sensitivity analyses
- Multi pollutant models

**Table S4.** Adjusted Pooled Effect Estimates for Parkinson's Disease Incidence by Geographical Region

| Geographical Region, country (n) | Pollutant | # Effect Estimates | Pooled Effect Estimate (95% CI) | $I^2$, p-value* | Prediction Interval |
|---|---|---|---|---|---|
| **Asia** China (2), South Korea (1) | $PM_{2.5}$ | 3 | 1.16 (0.96, 1.41) per 5 µg/$m^3$ increase | 43%, $X^2$=3.49, p = 0.17 | 0.46, 2.96 |
| **Europe** Finland (1), Italy (1), Netherlands (1) | $PM_{2.5}$ | 3 | 0.98 (0.93, 1.02) per 5 µg/$m^3$ increase | 0%, $X^2$=0.89, p = 0.64 | 0.80, 1.18 |
| **North America** Canada (3), USA (5) | $PM_{2.5}$ | 8 | 1.07 (0.98, 1.17) per 5 µg/$m^3$ increase | 85%, $X^2$= 52.86, p < 0.01 | 0.94, 1.14 |
| **North America** USA (3) | $PM_{10}$ | 3 | 0.99 (0.95, 1.03) per 15 µg/$m^3$ increase | 0%, $X^2$=0.83, p = 0.66 | 0.82, 1.19 |
| **Asia** Taiwan (2), South Korea (1), China (2) | $PM_{10}$ | 5 | 1.05 (0.83, 1.31) per 15 µg/$m^3$ increase | 78%, $X^2$= 18.60, p < 0.01 | 0.64, 1.71 |
| **Europe** Finland (1), Italy (1), Netherlands (1) | $PM_{10}$ | 3 | 0.98 (0.95, 1.00) per 15 µg/$m^3$ increase | 0%, $X^2$= 0.33, p = 0.85 | 0.78, 1.21 |
| **Asia** Taiwan (1), China (2) | $NO_2$ | 3 | 1.07 (0.99, 1.16) per 10 µg/$m^3$ increase | 0%, $X^2$= 0.34, p = 0.84 | 0.60, 1.90 |
| **North America** Canada (3), USA (1), | $NO_2$ | 4 | 1.02 (0.98, 1.06) per 10 µg/$m^3$ increase | 66%, $X^2$= 8.75, p = 0.03 | 0.94, 1.10 |
| **Europe** Denmark (1), Italy (1), Netherlands (1) | $NO_2$ | 3 | 1.05 (0.62, 1.79) per 10 µg/$m^3$ increase | 80%, $X^2$= 9.82, p < 0.01 | 0.08, 14.68 |
| **Europe** Denmark (1), Italy (1), Netherlands (1) | $NO_x$ | 3 | 1.03 (0.79, 1.35) per 17 µg/$m^3$ increase | 80%, $X^2$= 10.24, p < 0.01 | 0.22, 4.80 |

*p-value is from Cochran's Q (chi-squared) test for heterogeneity

**Table S5.** Adjusted Pooled Effect Estimates for Parkinson's Disease Incidence in Cohort Studies only

| Pollutant | # Effect Estimates | Pooled Effect Estimate | $I^2$, p-value* | Prediction Interval |
|---|---|---|---|---|
| $PM_{2.5}$ | 11 | 1.05 (0.99, 1.11) per 5 μg/$m^3$ increase | 80%, $X^2$= 49.97, p < 0.01 | 0.94, 1.17 |
| $PM_{10}$ | 6 | 0.98 (0.90, 1.08) per 15 μg/$m^3$ increase | 49%, $X^2$= 9.87, p = 0.08 | 0.86, 1.13 |
| $PM_{2.5-10}$ | 3 | 0.99 (0.97, 1.01) per 3 μg/$m^3$ increase | 39%, $X^2$= 3.26, p = 0.20 | 0.88, 1.11 |
| $NO_2$ | 6 | 1.01 (0.98, 1.05) per 10 μg/$m^3$ increase | 79%, $X^2$= 23.55, p < 0.01 | 0.95, 1.08 |
| Annual $O_3$ | 3 | 1.02 (0.96, 1.09) per 12 μg/$m^3$ increase | 0%, $X^2$= 1.82, p = 0.40 | 0.85, 1.24 |

*p-value is from Cochran's Q (chi-squared) test for heterogeneity

**Table S6.** Pooled Effect Estimates Overall, and by Study Risk Estimate Type (HR, OR) for Parkinson's Disease Incidence

| Pollutant | Overall Pooled Effect Estimate (95% CI) | # of studies | HR Only (95% CI) | # of HR studies | OR Only (95% CI) | # of OR studies |
|---|---|---|---|---|---|---|
| $PM_{2.5}$ | 1.06 (1.00, 1.12) per 5 μg/$m^3$ increase | 16 | 1.05 (0.98, 1.13) per 5 μg/$m^3$ increase | 8 | 1.13 (0.93, 1.36) per 5 μg/$m^3$ increase | 7 |
| $PM_{10}$ | 1.00 (0.93, 1.07) per 15 μg/$m^3$ increase | 12 | 0.99 (0.87, 1.12) per 15 μg/$m^3$ increase | 5 | 1.03 (0.89, 1.20) per 15 μg/$m^3$ increase | 6 |
| $PM_{2.5-10}$ | 0.99 (0.97, 1.01) per 3 μg/$m^3$ increase | 4 | Too Few | 2 | Too Few | 1 |
| $NO_2$ | 1.01 (0.97, 1.05) per 10 μg/$m^3$ increase | 11 | 1.01 (0.98, 1.05) per 10 μg/$m^3$ increase | 6 | 1.01 (0.83, 1.23) per 10 μg/$m^3$ increase | 5 |
| $NO_x$ | 1.01 (0.94, 1.08) per 17 μg/$m^3$ increase | 5 | Too Few | 1 | 1.03 (0.93, 1.15) per 17 μg/$m^3$ increase | 4 |
| CO | 1.08 (0.80, 1.47) per 0.2 mg/$m^3$ increase | 4 | NA | 0 | Same As Pooled | 4 |
| Annual $O_3$ | 1.01 (0.94, 1.09) per 12 μg/$m^3$ increase | 6 | Too Few | 1 | 1.04 (0.86, 1.25) per 12 μg/$m^3$ increase | 5 |
| Warm Season $O_3$ | 1.09 (0.39, 3.09) per 12 μg/$m^3$ increase | 3 | Too Few | 1 | Too Few | 2 |
| $SO_2$ | 1.09 (0.78, 1.53) per 3 μg/$m^3$ increase | 3 | NA | 0 | Same As Pooled | 3 |

*Note: Kirrane 2015 is counted as 2 studies since one estimate given for Iowa, another given for North Carolina*

**Table S7:** Overall Certainty of Evidence.

| | Factor | Present |
|---|---|---|
| **Factors Decreasing Confidence** | **Risk of bias (RoB):** The RoB assessment showed that only 21 out of 32 papers had 'probably high risk of bias' in one or more domains. When excluded in sensitivity analyses, there was a small, non-statistically significant effect on effect estimates that did not affect direction. All the other papers were assessed as 'probably low risk of bias' or 'definitely low risk of bias' in all domains. | No |
| | **Unexplained inconsistency:** There is unexplained inconsistency for all pollutant exposures and outcome pairs (e.g., PD and MS incidence) in the meta-analysis. | Yes |
| | **Indirectness:** All studies included were human studies where the exposure preceded the outcome. Studies address the relevant population of interest, with the exposure of interest, and the overall effect that is intended to be measured is true and relevant to the real-world context. Included studies had a long follow period between exposure and outcome ascertainment (e.g., at least more than one year). | No |
| | **Imprecision:** All but one study fulfilled the 'not serious' criteria based on ratio (≤10) and absolute difference (≤100) of upper to lower 95% confidence interval values. | No |
| | **Publication bias:** The results of our Egger's linear regression test demonstrate the presence of funnel plot asymmetry for $PM_{2.5}$ and $PM_{10}$ exposure for PD, but not $NO_2$. | Yes |
| **Factors Increasing Confidence** | **Large magnitude of effect:** What defines a large magnitude of effect in an environmental context is a debated topic. None of the pooled effect estimates were above 1.1. | No |
| | **Dose-response relationship:** Four or more studies reported categorical results for $NO_2$, $PM_{2.5}$ and $PM_{10}$ with PD as the outcome. These have been included in the visualisations of the ERFs and did not show a clear dose-response relationship, with both monotonic and non-monotonic relationships (Figure S3).<br><br>ERF's published in primary papers did show some dose-response relationships. For $NO_2$, Ritz et al (2016) showed a linear monotonic relationship and Sungyang et al (2021) a non-monotonic relationship. For $PM_{2.5}$, most of the primary papers reported the highest category having high effect estimates than the reference, but only two showed incremental increases (Lee et al 2022 and Kwon et al 2023). For $PM_{10}$, Chen et al 2017a reported a linear monotonic relationship between pollutant concentration and effect estimate, but four out ten papers reported the effect estimate for the highest category was lower than the reference effect estimate. | No |

| | | |
|---|---|---|
| | Exposure-outcome pairs for MS and MND did not meet the threshold for the creation of ERFs due to an insufficient number of papers with unique categorical data that represented at least three categories of exposure. | |
| | **Residual confounding factors:** Residual confounding in the relationship between air pollution and outcomes of PD, MS and MND may emerge from several sources. This includes occupational exposures, and genetic factors. All studies included in the meta-analysis adjusted for different confounders in their analysis, but there is still a likelihood that residual confounding may exist within our meta-analysis. We conducted a meta-analysis with unadjusted effect estimates, but only had enough studies for $PM_{10}$ and $PM_{2.5}$ exposure and PD as the outcome. The adjusted and unadjusted pooled effect estimates differed by 0.02 (higher for adjusted over unadjusted for both pollutants). Covariates included in the adjustment may not significantly impact the overall effects, but as it was only possible to assess this for two pollutant-outcome pairs, this factor cannot be viewed as increasing confidence. | No |
| | **Cross population and cross-study-design consistency:** Whilst there were consistent results across geographical regions and study types (e.g., cohort and case-control), none of subgroup analyses reached statistical significance. | Yes |

**Table S8.** Pooled effect estimates per continuous exposure by outcome, pollutant, and adjustment stratified by whether study with largest weight (smallest SE) was excluded or not.

| Outcome | Adjusted | Pollutant | Removed | N | Pooled effect estimate | 95% CI | PI | $Tau^2$ | $I^2$ |
|---|---|---|---|---|---|---|---|---|---|
| PD | No | $PM_{2.5}$ | - | 4 | 1.044 | 0.846-1.287 | 0.7-1.556 | 0.006 | 0.838 |
| PD | No | $PM_{2.5}$ | Rumrich et al. (2023) | 3 | 1.08 | 0.683-1.709 | 0.062-18.832 | 0.034 | 0.892 |
| PD | Yes | CO | - | 4 | 1.082 | 0.796-1.472 | 0.684-1.712 | 0.008 | 0.829 |
| PD | Yes | CO | Lee et al. (2016) | 3 | 1.165 | 0.46-2.953 | 0.089-15.247 | 0.027 | 0.862 |
| PD | Yes | $NO_2$ | - | 11 | 1.011 | 0.97-1.053 | 0.944-1.082 | 0.001 | 0.76 |
| PD | Yes | $NO_2$ | Shin et al. (2018) | 10 | 1.011 | 0.958-1.067 | 0.926-1.104 | 0.001 | 0.76 |
| PD | Yes | $NO_x$ | - | 5 | 1.008 | 0.942-1.079 | 0.898-1.132 | 0.001 | 0.819 |
| PD | Yes | $NO_x$ | Cerza et al. (2018) | 4 | 1.034 | 0.928-1.152 | 0.818-1.306 | 0.002 | 0.531 |
| PD | Yes | $PM_{10}$ | - | 12 | 1.001 | 0.934-1.072 | 0.885-1.133 | 0.003 | 0.713 |
| PD | Yes | $PM_{10}$ | Lee et al. (2016) | 11 | 1.018 | 0.942-1.1 | 0.893-1.16 | 0.003 | 0.608 |
| PD | Yes | $PM_{2.5}$ | - | 16 | 1.06 | 1.001-1.123 | 0.948-1.186 | 0.002 | 0.805 |
| PD | Yes | $PM_{2.5}$ | Palacios et al. (2017) | 15 | 1.08 | 1.011-1.154 | 0.94-1.241 | 0.004 | 0.765 |
| PD | Yes | $PM_{2.5-10}$ | - | 4 | 0.991 | 0.974-1.009 | 0.954-1.03 | 0 | 0.28 |
| PD | Yes | $PM_{2.5-10}$ | Palacios et al. (2017) | 3 | 0.982 | 0.96-1.003 | 0.89-1.082 | 0 | 0 |
| PD | Yes | annual $O_3$ | - | 6 | 1.012 | 0.938-1.093 | 0.89-1.151 | 0.001 | 0.52 |
| PD | Yes | annual $O_3$ | Cerza et al. (2018) | 5 | 1.04 | 0.864-1.252 | 0.722-1.499 | 0.009 | 0.409 |

**Table S9.** Pooled effect estimates per continuous exposure by outcome, pollutant, and adjustment stratified by whether study was assigned low risk of bias (i.e. no domains with "Probably high risk of bias") or high risk of bias (i.e., at least 1 domain with "Probably high risk of bias").

| Outcome | Adjusted | Pollutant | Bias | K | Pooled effect estimate | 95% CI | PI | $Tau^2$ | $I^2$ |
|---|---|---|---|---|---|---|---|---|---|
| PD | No | $PM_{10}$ | low | 3 | 0.9826 | 0.9466-1.0199 | 0.7804-1.2372 | 0 | 0 |
| PD | No | $PM_{2.5}$ | low | 4 | 1.0436 | 0.8464-1.2868 | 0.7001-1.5558 | 0.0062 | 0.8375 |
| PD | Yes | $NO_2$ | high | 6 | 1.0147 | 0.9600-1.0724 | 0.9387-1.0968 | 6.00E-04 | 0.7398 |
| PD | Yes | $NO_2$ | low | 5 | 1.0294 | 0.8907-1.1897 | 0.8230-1.2875 | 0.0035 | 0.6673 |
| PD | Yes | $NO_x$ | low | 3 | 1.0285 | 0.7861-1.3456 | 0.2206-4.7955 | 0.0099 | 0.8047 |
| PD | Yes | $PM_{10}$ | high | 7 | 1.0122 | 0.8831-1.1603 | 0.8359-1.2258 | 0.0043 | 0.8067 |
| PD | Yes | $PM_{10}$ | low | 5 | 0.9897 | 0.9243-1.0597 | 0.8901-1.1005 | 6.00E-04 | 0.256 |
| PD | Yes | $PM_{2.5}$ | high | 10 | 1.0534 | 1.0000-1.1096 | 0.9541-1.1630 | 0.0015 | 0.7523 |
| PD | Yes | $PM_{2.5}$ | low | 6 | 1.0927 | 0.9185-1.2999 | 0.7881-1.5149 | 0.0112 | 0.8762 |
| PD | Yes | $SO_2$ | high | 3 | 1.0913 | 0.7779-1.5309 | 0.0767-15.525 | 0.0316 | 0.9894 |
| PD | Yes | annual $O_3$ | high | 5 | 1.0402 | 0.8645-1.2516 | 0.7220-1.4987 | 0.0086 | 0.409 |
| PD | Yes | warm season $O_3$ | high | 3 | 1.0915 | 0.3859-3.0870 | 0.0128-93.1528 | 0.0775 | 0.5392 |

**Table S10.** Adjusted Pooled Effect Estimates for Parkinson's Disease Incidence in Sensitivity Analysis Excluding Studies from our Meta-Analysis that Adjusted for Comorbidities.

| Pollutant | # Effect Estimates | Pooled Effect Estimate | $I^2$, p-value* | Prediction Interval |
|---|---|---|---|---|
| $PM_{2.5}$ | 13 | 1.08 (1.00, 1.16) per 5 $\mu g/m^3$ increase | 80%, $X^2$= 59.35, p < 0.05 | 0.93, 1.26 |
| $PM_{10}$ | 10 | 0.99 (0.91, 1.08) per 15 $\mu g/m^3$ increase | 72%, $X^2$= 31.68, p < 0.05 | 0.87, 1.14 |
| $PM_{2.5\text{-}10}$ | 4 | 0.99 (0.97, 1.01) per 3 $\mu g/m^3$ increase | 28%, $X^2$= 4.17, p = 0.24 | 0.95, 1.03 |
| $NO_2$ | 8 | 1.00 (0.93, 1.07) per 10 $\mu g/m^3$ increase | 77%, $X^2$= 30.28, p < 0.01 | 0.91, 1.10 |
| $NO_x$ | 4 | 1.01 (0.92, 1.11) per 17 $\mu g/m^3$ increase | 86%, $X^2$= 22.11, p < 0.01 | 0.86, 1.19 |
| CO | 3 | 1.15 (0.53, 2.51) per 0.2 $mg/m^3$ increase | 88%, $X^2$= 17.00, p < 0.01 | 0.14, 9.55 |
| Annual $O_3$ | 5 | 1.01 (0.92, 1.11) per 12 $\mu g/m^3$ increase | 61%, $X^2$= 10.16, p = 0.04 | 0.86, 1.19 |
| Warm Season $O_3$ | 3 | 1.09 (0.39, 3.09) per 12 $\mu g/m^3$ increase | 54%, $X^2$= 4.34, p = 0.11 | 0.01, 93.15 |

*p-value is from Cochran's Q (chi-squared) test for heterogeneity

**Table S11.** Adjusted Pooled Effect Estimates for Parkinson's Disease Incidence in Sensitivity Analysis Excluding Studies from our Meta-Analysis that Adjusted for Cardiovascular Disease.

| Pollutant | # Effect Estimates | Pooled Effect Estimate | $I^2$, p-value* | Prediction Interval |
|---|---|---|---|---|
| $PM_{2.5}$ | 14 | 1.06 (0.99, 1.13) per 5 µg/$m^3$ increase | 78%, $X^2$= 59.35, p < 0.05 | 0.93, 1.21 |
| $PM_{10}$ | 11 | 0.99 (0.93, 1.06) per 15 µg/$m^3$ increase | 69%, $X^2$= 31.68, p < 0.05 | 0.88, 1.11 |
| $PM_{2.5\text{-}10}$ | 4 | 0.99 (0.97, 1.01) per 3 µg/$m^3$ increase | 28%, $X^2$= 4.17, p = 0.24 | 0.95, 1.03 |
| $NO_2$ | 8 | 1.00 (0.93, 1.07) per 10 µg/$m^3$ increase | 77%, $X^2$= 30.28, p < 0.01 | 0.91, 1.10 |
| $NO_x$ | 4 | 1.01 (0.92, 1.11) per 17 µg/$m^3$ increase | 86%, $X^2$= 22.11, p < 0.01 | 0.86, 1.19 |
| CO | 3 | 1.15 (0.53, 2.51) per 0.2 mg/$m^3$ increase | 88%, $X^2$= 17.00, p < 0.01 | 0.14, 9.55 |
| Annual $O_3$ | 5 | 1.01 (0.92, 1.11) per 12 µg/$m^3$ increase | 61%, $X^2$= 10.16, p = 0.04 | 0.86, 1.19 |
| Warm Season $O_3$ | 3 | 1.09 (0.39, 3.09) per 12 µg/$m^3$ increase | 54%, $X^2$= 4.34, p = 0.11 | 0.01, 93.15 |

*p-value is from Cochran's Q (chi-squared) test for heterogeneity

**Figure S1**. Forest plot of subgroup meta-analysis by Exposure Assessment methods for risk of Parkinson's Disease Incidence from $PM_{2.5}$ exposure.

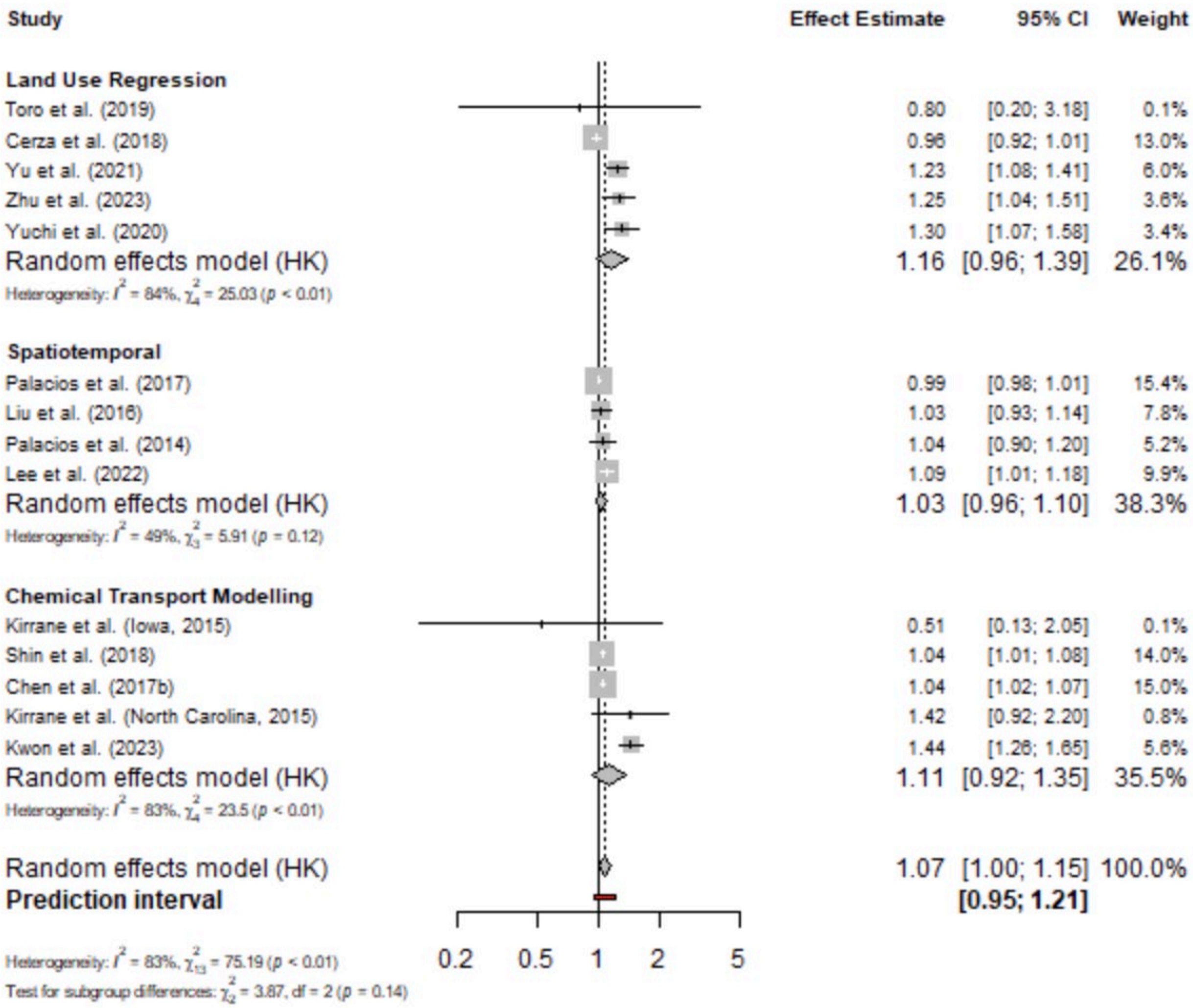

**Figure S2**. Forest plot of subgroup meta-analysis by Exposure Assessment methods for risk of Parkinson's Disease Incidence from $PM_{10}$ exposure.

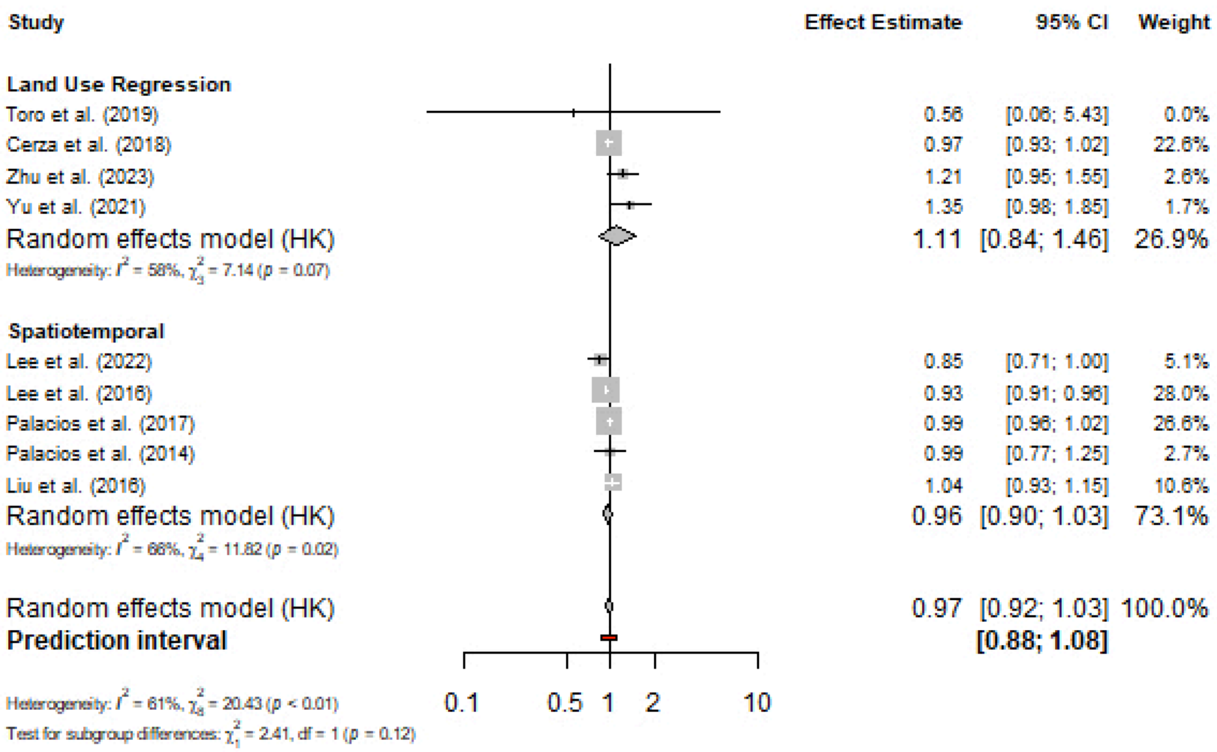

**Figure S3**. ERF curves for categorical estimates.

a) ERF curve for $PM_{2.5}$ estimates.

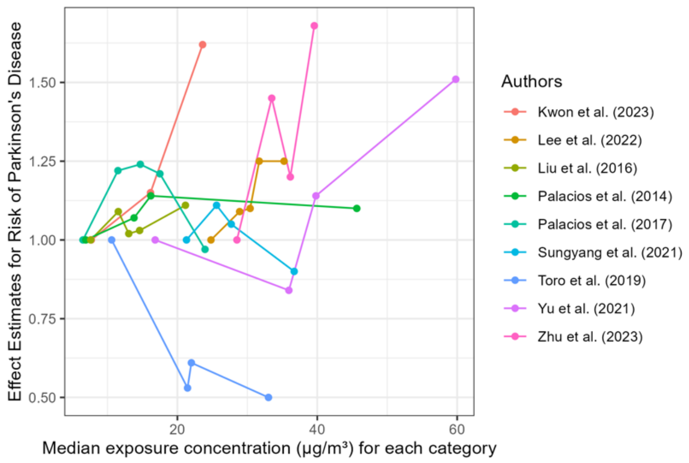


b) ERF curve for $PM_{10}$ estimates.

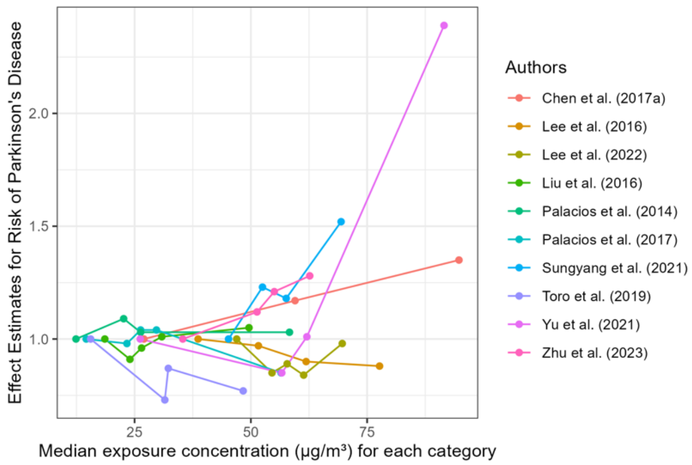


c) ERF curve for $NO_2$ estimates.

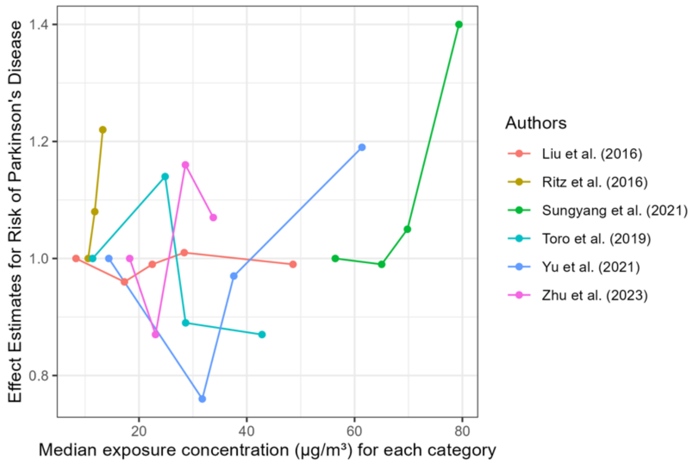

Supplementary Material:

S1: List of excluded studies and reason for exclusion at full text screening stage

https://docs.google.com/spreadsheets/d/1X06zVqINJ7QZLZ7JMm7AnNCZDqeNh2KgSyWiyuqx4fE/edit?gid=1052957813#gid=1052957813

S2: Data Extraction Form

https://docs.google.com/spreadsheets/d/1kmv5NwF8vATMpBir_5ZVVkCr7BWZq60N/edit?gid=1016733053#gid=1016733053

S3: Complete table of extracted data points

https://docs.google.com/spreadsheets/d/1E98kaFxheJ1K5WpCAcYwjgMuVUB7-HrmZdfvXc8WJtg/edit?usp=sharing
